# Can drinking water, sanitation, handwashing, and nutritional interventions reduce antibiotic use in young children?

**DOI:** 10.1101/2024.05.10.24307069

**Authors:** Ayse Ercumen, Andrew N. Mertens, Zachary Butzin-Dozier, Da Kyung Jung, Shahjahan Ali, Beryl S. Achando, Gouthami Rao, Caitlin Hemlock, Amy J. Pickering, Christine P. Stewart, Sophia T. Tan, Jessica A. Grembi, Jade Benjamin-Chung, Marlene Wolfe, Gene G. Ho, Md. Ziaur Rahman, Charles D. Arnold, Holly N. Dentz, Sammy M Njenga, Dorie Meerkerk, Belinda Chen, Maya Nadimpalli, Mohammad Aminul Islam, Alan E. Hubbard, Clair Null, Leanne Unicomb, Mahbubur Rahman, John M. Colford, Stephen P. Luby, Benjamin F. Arnold, Audrie Lin

## Abstract

Frequent antibiotic use in areas with high infection burdens can lead to antimicrobial resistance and microbiome alterations. Reducing pathogen exposure and child undernutrition can reduce infections and subsequent antibiotic use. We assessed effects of water, sanitation, handwashing (WSH) and nutrition interventions on pediatric antibiotic use in Bangladesh and Kenya, using longitudinal data from birth cohorts (at ages 3-28 months) enrolled in the WASH Benefits cluster-randomized trials. Over 50% of children used antibiotics at least once in the last 90 days. In Bangladesh, the prevalence of using antibiotics at least once was 10-14% lower in groups receiving WSH (prevalence ratio [PR]=0.90 (0.82-0.99)), nutrition (PR=0.86 (0.78-0.94)), and nutrition+WSH (PR=0.86 (0.79-0.93)) interventions. The prevalence of using antibiotics multiple times was 24-35% lower in intervention arms. Intervention effects were strongest when the birth cohort was younger. In Kenya, interventions did not affect antibiotic use. Improving WSH and nutrition can reduce antibiotic use in some low- and middle-income settings. Studies should assess whether such reductions translate to reduced carriage of antimicrobial resistance.

## Introduction

Antimicrobial resistance (AMR) was associated with an estimated 4.95 million deaths in 2019 ^1^, most in low-and-middle income countries (LMICs). Community carriage of antimicrobial resistant bacteria ^2^ and their abundance in sewage ^3^ is higher in LMICs – especially in Sub-Saharan Africa and South Asia – than high-income countries. Reasons may include densely populated conditions, lack of safe drinking water and sanitation ^4^, and widespread availability and frequent use of antibiotics providing selective advantages for resistant strains ^5^. A longitudinal study following birth cohorts in eight countries in South America, sub-Saharan Africa and Asia found the highest pediatric antibiotic use in South Asia ^6^. In Bangladesh, 98% of the children in the study used antibiotics before the age of 6 months and had an average of 10 courses of antibiotics per child-year ^6^. Similarly, in Kenya, children have an average of 22 antibiotic prescriptions between birth and age of 5 years ^7^. In comparison, a child in the US takes <2 courses of antibiotics per child-year ^8^. Primary reasons for antibiotic usage in LMICs are diarrheal and respiratory infections, which are associated with a high burden of childhood morbidity and mortality ^6,7^.

Children are susceptible to enteric and respiratory infections when they are frequently exposed to pathogens due to poor water, sanitation, and hygiene (WASH) conditions. As of 2020, 30% of sub-Saharan Africa (and 13% of rural sub-Saharan Africa) had access to safely managed water services ^9^. Similarly, 21% of sub-Saharan Africa and 47% of South Asia had access to safely managed sanitation in 2020 ^9^. Poor WASH conditions are estimated to account for 62% of deaths from diarrhea among children <5 years globally ^10^. Early life antibiotic use (e.g., to treat childhood diarrheal infections) can in turn further increase the risk of diarrhea ^11,12^. Additionally, repeated episodes of diarrhea can lead to malnutrition, and immune systems weakened by poor nutritional status put children at risk of further infections ^13^. Improving WASH conditions in LMICs may reduce antibiotic use by decreasing incidences of childhood diarrhea ^14^ and respiratory infections ^15,16^. Similarly, improved nutrition can make children less susceptible to infections ^17^ and reduce subsequent antibiotic use. A modeling study estimated that universal access to improved water and sanitation could reduce antibiotic use by 60% ^18^. However, there are scarce empirical data on the effect of WASH and nutrition interventions on antibiotic use by children. A single randomized controlled trial to date found that community-level water chlorination reduced reported antibiotic use by 7% among children in urban Bangladesh, along with a 23% reduction in diarrhea ^19^.

Here, we utilize data from two cluster-randomized controlled trials of individual and combined water, sanitation, handwashing (WSH) and nutrition interventions in rural Bangladesh and Kenya. In Bangladesh, children receiving WSH and nutrition interventions had reduced prevalence of diarrhea ^20^ and respiratory infections ^21^ compared to controls. In Kenya, children receiving the nutrition intervention had marginally lower prevalence of respiratory infections than controls; there were no other intervention effects ^22,23^. In both countries, the nutrition intervention improved child linear growth ^20,22^. It is plausible that children that experienced fewer infections or better growth consequently used less antibiotics during the trial. The objective of the current study was to assess whether these interventions reduced antibiotic use in children in rural Bangladesh and Kenya. For each country, we assessed the effects of the WSH, nutrition and nutrition plus WSH (N+WSH) interventions compared to controls, and the effects of the combined N+WSH intervention compared to WSH and nutrition interventions alone, with subgroup analyses by child age and sex. We followed CONSORT guidelines for cluster-randomized trials (see Supplemental Table S1 for CONSORT checklist).

## Results and discussion

### Enrollment

In Bangladesh, 5551 pregnant women in 720 clusters were enrolled between 31 May 2012 and 7 July 2013. Antibiotic use was recorded among children in the birth cohort participating in a longitudinal substudy conducted to assess environmental enteric dysfunction (EED), which included 1131 children at age 3 months, 1531 children at 14 months and 1531 children at 28 months (Supplemental Fig S1). Of these, antibiotic data were available for 1102 children (97%) at 3 months and 1528 children (>99%) each at 14 and 28 months (Supplemental Fig S1). In Kenya, 8246 pregnant women in 702 clusters were enrolled between 27 November 2012 and 21 May 2014. The EED substudy included 1493 children at age 6 months, 1504 children at 17 months and 1444 children at 22 months (Supplemental Fig S2). Antibiotic data were available for 1438 children (96%) at 6 months, 1449 children (96%) at 17 months and 1393 children (97%) at 22 months (Supplemental Fig S2). Among households enrolled in the EED substudy, characteristics were balanced across study arms and similar to the households enrolled in the full trial for both countries (Supplemental Tables S2, S3). Children lost to follow-up at the latter two measurement points were similar in their characteristics to those that completed all three follow-ups (Supplemental Tables S4, S5).

### Antibiotic use

In Bangladesh, 63% (635) of children in the control group used antibiotics at least once and 25% (248) multiple times in the last 90 days, for a mean of 5 total days (standard deviation [SD]=6) (Table 1). Use was highest at the 14-month measurement, where 75% of children in the control group used antibiotics in the last 90 days, and use was somewhat higher for boys than girls (Supplemental Table S6). Caregivers reported 24 distinct antibiotics. The most common antibiotic classes were penicillins (34.4%), cephalosporins (30.6%) and macrolides (22.5%), while the most common antibiotics were amoxycillin (32.1%), azithromycin (19.6%) and cefixime (12.3%).

**Table 1.** Caregiver-reported antibiotic use in last 3 months among young children enrolled in the control group of the WASH Benefits Bangladesh and Kenya trials.

|  | Bangladesh |  | Kenya |  |
| --- | --- | --- | --- | --- |
|  | N | % (n) / mean (SD) | N | % (n) / mean (SD) |
| Used antibiotics $\geq 1$ time | 1005 | 63.2 (635) | 1143 | 52.6 (601) |
| Used antibiotics $\geq 2$ times | 1005 | 24.7 (248) | 1143 | 13.3 (152) |
| Episodes of antibiotic use | 1005 | 0.98 (0.98) | 1143 | 0.68 (0.75) |
| Total days of antibiotic use | 995 | 5.06 (5.81) | 1118 | 3.95 (12.1) |

SD: Standard deviation

In Kenya, 53% (601) of children in the control group used antibiotics at least once and 13% (152) multiple times in the last 90 days, for a mean of 4 total days (SD=12) (Table 1). Use appeared highest at the 6-month measurement and similar for girls and boys (Supplemental Table S7). Caregivers reported 15 distinct antibiotics. The most common classes were sulfonamides (52.6%), penicillins (39.6%) and nitroimidazoles (7.2%), and the most common antibiotics were cotrimoxazole (52.6%), amoxycillin (38.7) and metronidazole (7.2%).

### Intervention effects

All interventions reduced antibiotic use in Bangladesh compared to controls (Fig 1, Fig 2). The percentage of children who used antibiotics at least once in the last 90 days was 10-14% lower among children receiving any intervention than controls (WSH prevalence ratio [PR]=0.90, 95% CI: 0.82-0.99, p=0.03; nutrition PR=0.86, 95% CI: 0.78-0.94, p<0.001, N+WSH PR=0.86, 95% CI: 0.79-0.93, p<0.001, Fig 1, Supplemental Table S8). The percent of children who used antibiotics multiple times in the last 90 days was 26-35% lower among children receiving interventions than controls (WSH PR=0.74, 95% CI: 0.63-0.87, p<0.001, nutrition PR=0.66, 95% CI: 0.56-0.79, p<0.001; N+WSH PR=0.65, 95% CI: 0.55-0.78, p<0.001, Fig 1, Supplemental Table S8). In all intervention arms, episodes of antibiotic use was reduced by 0.17-0.21 episodes and total days of antibiotic use by approximately 1 day compared to controls (Fig 2, Supplemental Table S8). The N+WSH intervention did not additionally reduce antibiotic use compared to the WSH and nutrition interventions (Fig 1, Fig 2, Supplemental Table S9). In Kenya, interventions were not associated with reduced antibiotic use compared to controls (Fig 1, Fig 2, Supplemental Table S10). In the N+WSH arm, antibiotic use was similar to the WSH and nutrition arms (Fig 1, Fig 2, Supplemental Table S11).

**Fig 1.**
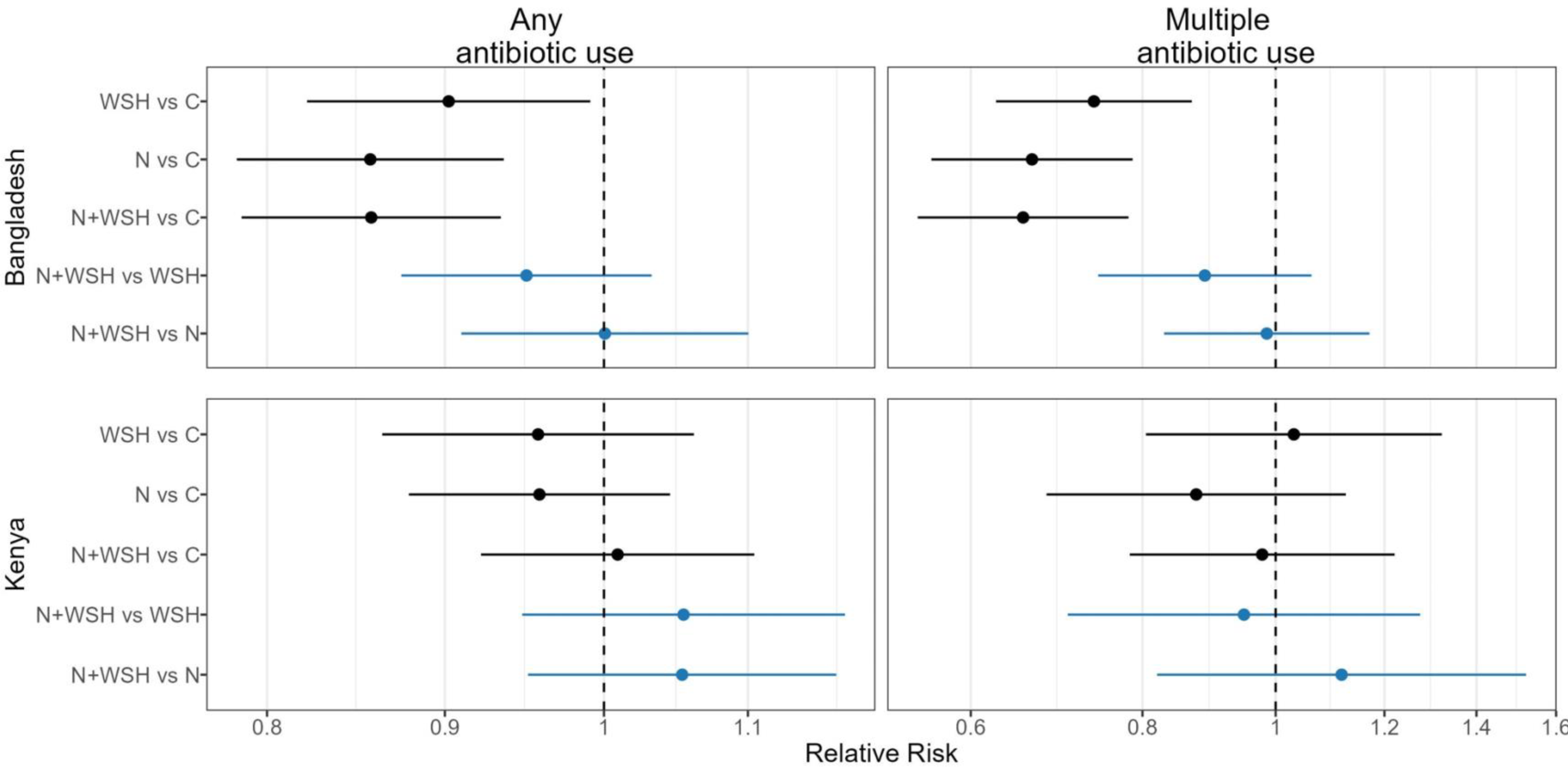
Relative effects of water, sanitation, handwashing (WSH), nutrition (N) and nutrition plus WSH (N+WSH) interventions on the prevalence of using antibiotics at least once and using antibiotics more than once in the last 3 months among young children in Bangladesh and Kenya. Estimates in black denote comparisons against the control (C) group who received no intervention. Estimates in blue denote comparisons of the N+WSH intervention group to the WSH and N intervention groups.

**Fig 2.**
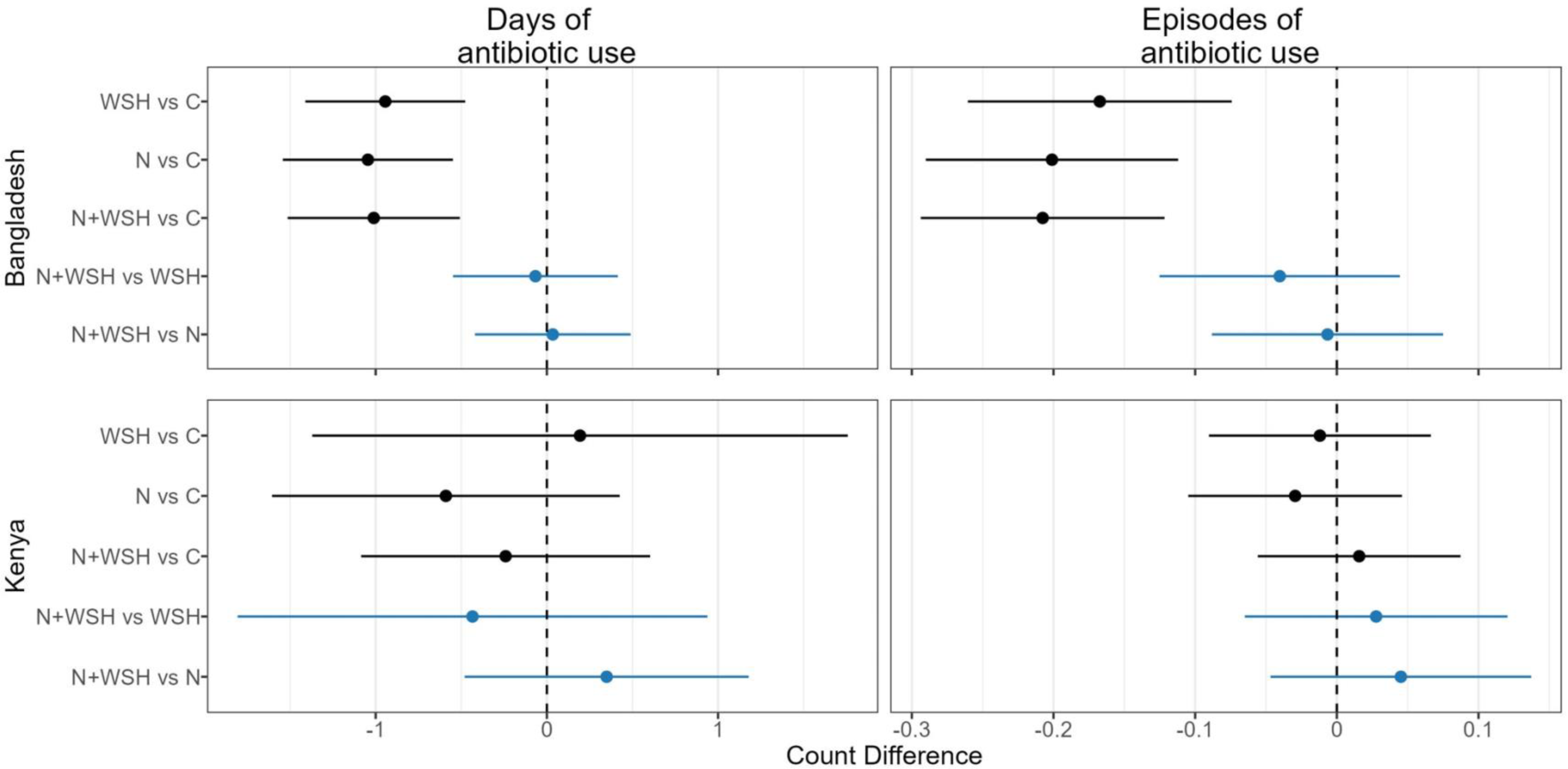
Absolute effects of water, sanitation, handwashing (WSH), nutrition (N) and nutrition plus WSH (N+WSH) interventions on the number of days and episodes of antibiotic use in the last 3 months among young children in Bangladesh and Kenya. Estimates in black denote comparisons against the control (C) group who received no intervention. Estimates in blue denote comparisons of the N+WSH intervention group to the WSH and N intervention groups.

### Effect modification

In Bangladesh, most antibiotic use metrics showed effect modification by child age (interaction p-values<0.20, Supplemental Tables S12-S13) and intervention effects were strongest for the youngest children. Children at age 3 months in any intervention arm experienced reduced antibiotic use compared to controls (Fig 3, Fig 4). In this age group, the percent of children who used antibiotics at least once in 90 days was 24-33% lower in the nutrition and N+WSH arms (Supplemental Tables S12-S13). Notably, in this age group, the percent of children who used antibiotics multiple times was 43% lower in the WSH arm (PR=0.57, 95% CI: 0.37-0.86), 49% lower in the nutrition arm (PR=0.51, 95% CI: 0.33-0.78) and 52% lower in the N+WSH arm (PR=0.48, 95% CI: 0.33-0.70) compared to controls (Supplemental Tables S12-S13). In this age group, all interventions also reduced episodes of antibiotic use by 0.25-0.34 episodes and total days of antibiotic use by 1.23-1.52 days (Supplemental Table S13). At 14 months, interventions reduced the percent of children who used antibiotics multiple times by 23-30% and episodes of antibiotic use by 0.15-0.19 episodes (Supplemental Tables S12-S13). Days of antibiotic use was reduced by approximately 1 day in the WSH and nutrition arms but not N+WSH arm compared to controls (Supplemental Table S13). At 28 months, the WSH intervention had no effect on any metric of antibiotic use (Fig 3, Fig 4). The nutrition intervention reduced single antibiotic use by 15% and episodes of antibiotic use by 0.12 episodes, while the N+WSH intervention reduced multiple antibiotic use by 36%, episodes of antibiotic use by 0.15 episodes and days of antibiotic use by approximately 1 day (Supplemental Tables S12-S13).

**Fig 3.**
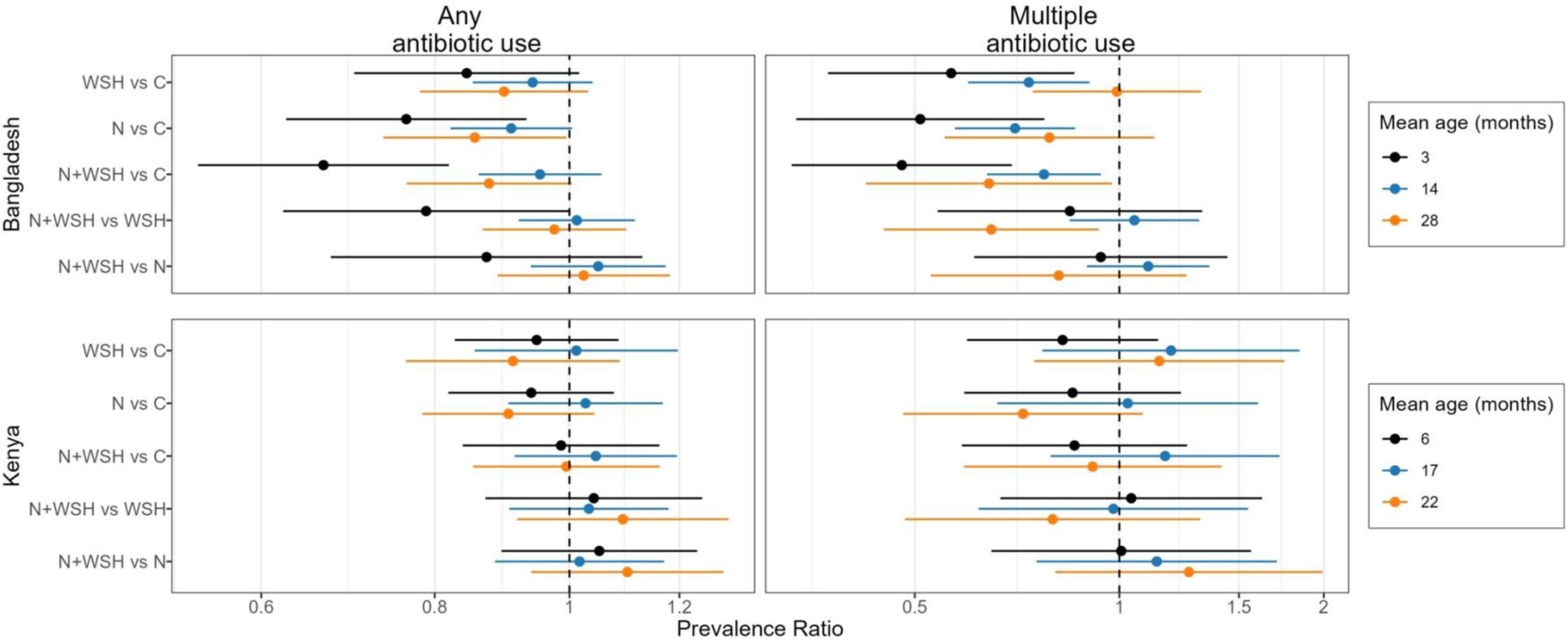
Subgroup analysis by mean child age for relative effects of water, sanitation, handwashing (WSH), nutrition (N) and nutrition plus WSH (N+WSH) interventions on the prevalence of using antibiotics at least once and using antibiotics more than once in the last 3 months among young children in Bangladesh and Kenya. The control group (C) received no intervention. Antibiotic use was recorded when the children were on average 3, 14 and 28 months old in Bangladesh, and 6, 17 and 22 months old in Kenya.

**Fig 4.**
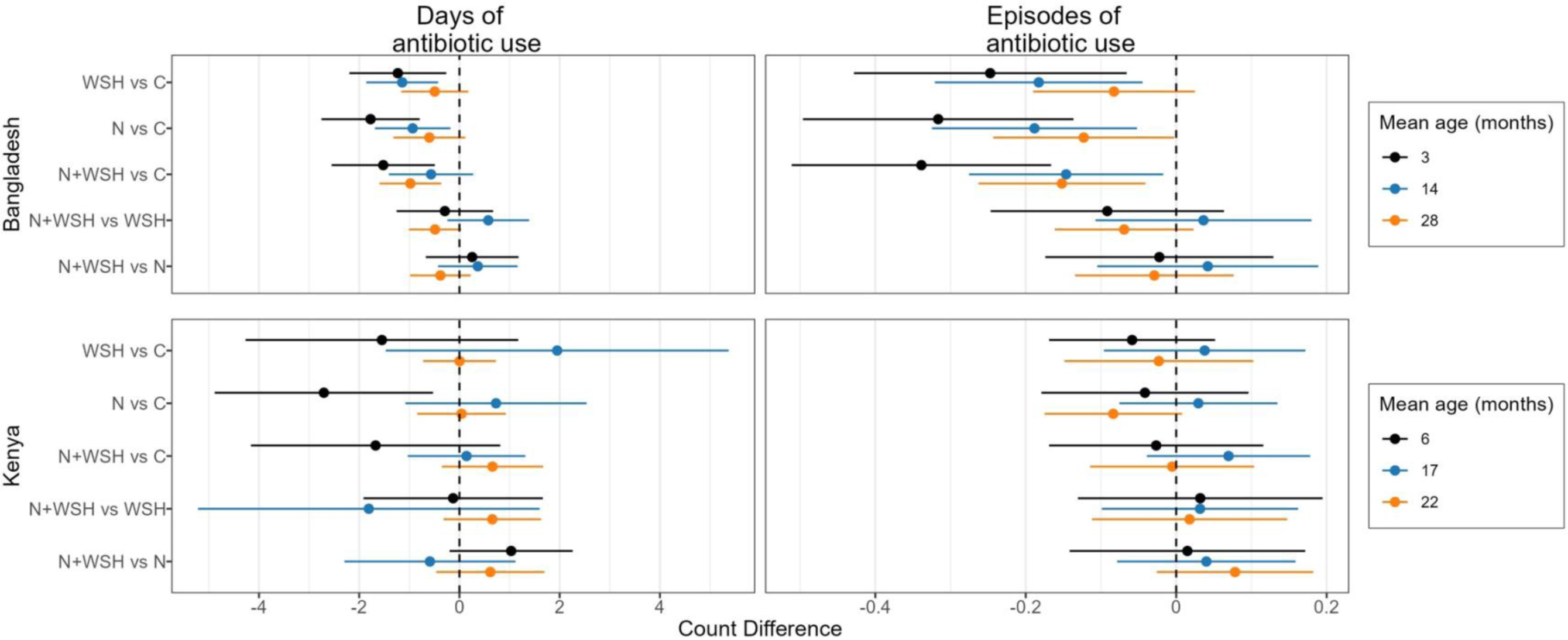
Subgroup analysis by mean child age for absolute effects of water, sanitation, handwashing (WSH), nutrition (N) and nutrition plus WSH (N+WSH) interventions on the number of days and episodes of antibiotic use in the last 3 months among young children in Bangladesh and Kenya. The control group (C) received no intervention. Antibiotic use was recorded when the children were on average 3, 14 and 28 months old in Bangladesh, and 6, 17 and 22 months old in Kenya.

In Bangladesh, use of antibiotics at least once in 90 days showed effect modification by child sex (interaction p-values<0.20, Supplemental Tables S12-S13); all three interventions reduced the percent of girls but not boys who used antibiotics at least once in 90 days. All interventions reduced the percent of both girls and boys who used antibiotics multiple times in 90 days and the episodes and days of antibiotic use (Fig 5, Fig 6, Supplemental Tables S12-S13). Overall, the N+WSH intervention was not any more effective than the WSH and nutrition interventions in subgroup analyses by age and sex (Figs 3-6, Supplemental Tables S12-S13).

**Fig 5.**
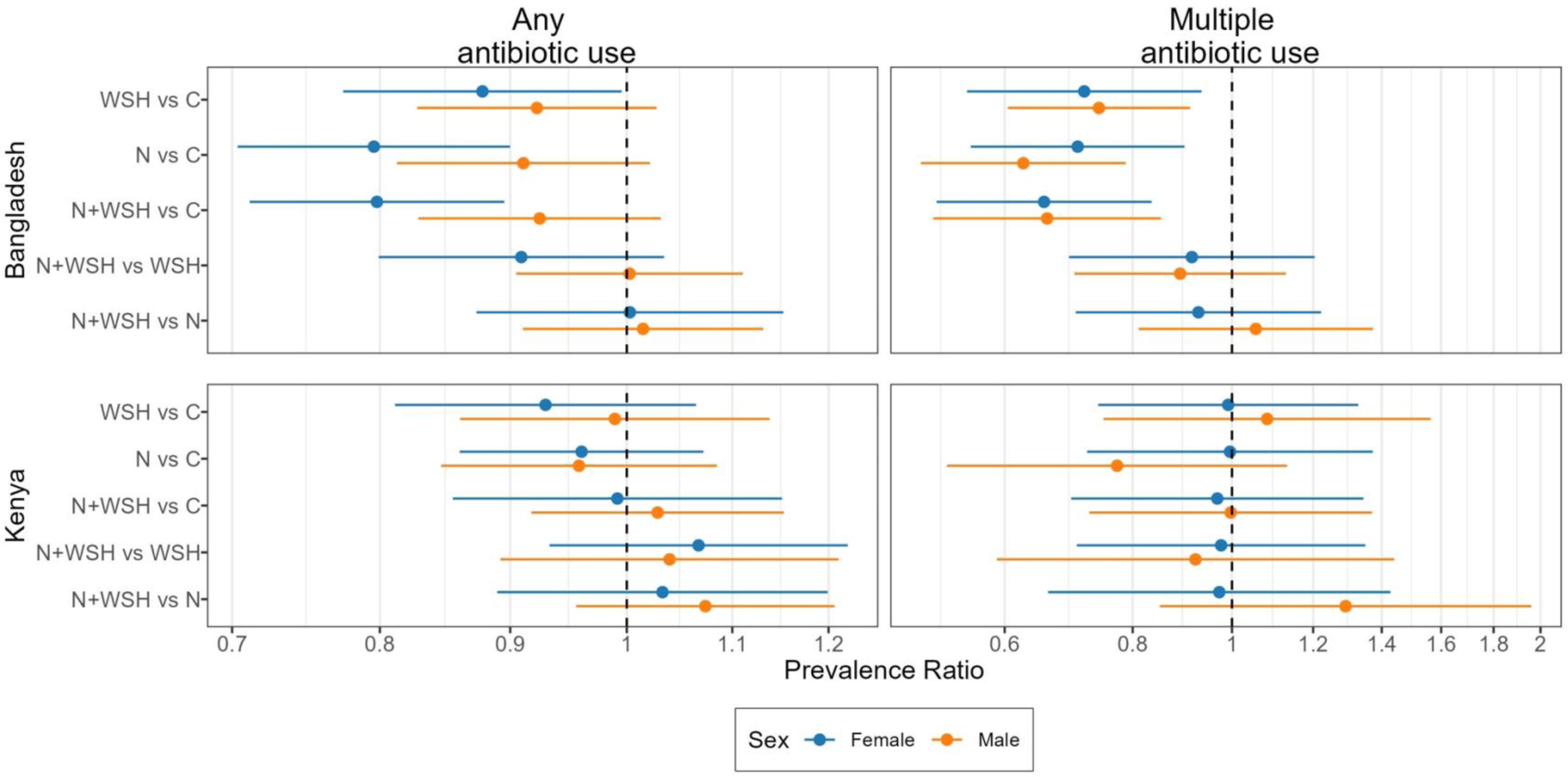
Subgroup analysis by child sex for relative effects of water, sanitation, handwashing (WSH), nutrition (N) and nutrition plus WSH (N+WSH) interventions on the prevalence of using antibiotics at least once and using antibiotics more than once in the last 3 months among young children in Bangladesh and Kenya. The control group (C) received no intervention.

**Fig 6.**
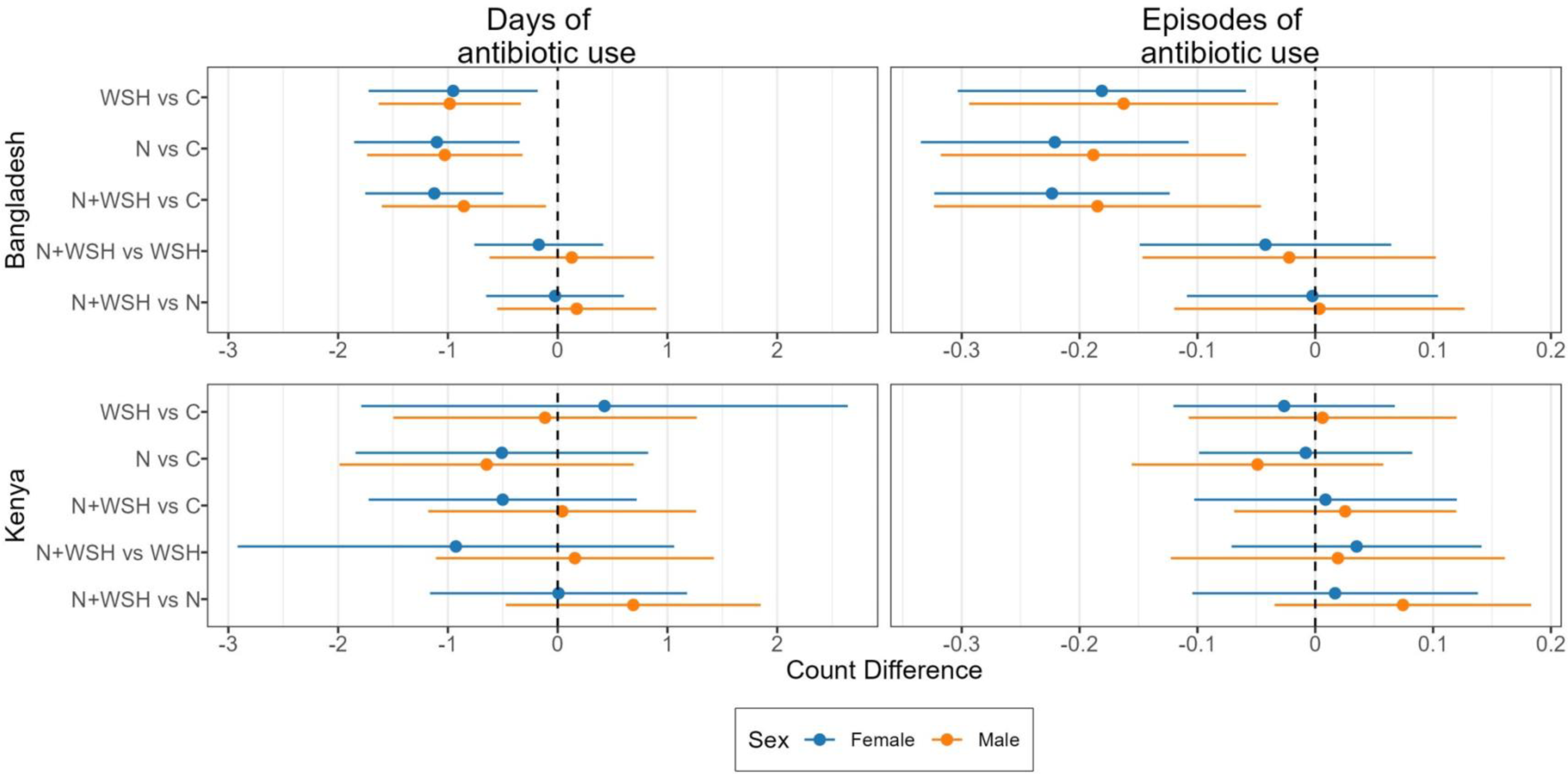
Subgroup analysis by child sex for absolute effects of water, sanitation, handwashing (WSH), nutrition (N) and nutrition plus WSH (N+WSH) interventions on the number of days and episodes of antibiotic use in the last 3 months among young children in Bangladesh and Kenya. The control group (C) received no intervention.

In Kenya, there was no consistent evidence of effect modification by child age or sex (Figs 3-6, Supplemental Tables S14-S15). At age 6 months, the nutrition intervention reduced total days of antibiotic use by approximately 3 days (count difference [CD]=-2.71, 95% CI: -4.88, -0.53) (Supplemental Table S15). There were no other intervention effects in any age or sex subgroup (Figs 3-6, Supplemental Tables S14-S15).

### Secondary and sensitivity analyses

Unadjusted and adjusted effect estimates were similar (Supplemental Tables S8-S11). In sensitivity analyses, all interventions in Bangladesh reduced the percent of children who used antibiotics at least once in the last month by 10-16% but had no effect on antibiotic use in the last two weeks (Supplemental Table S16).

## Discussion

More than half of children enrolled in both Bangladesh and Kenya used antibiotics at least once in the previous 90 days. Antibiotic use was reduced among children randomized to receive WSH, nutrition and nutrition+WSH interventions in Bangladesh but not in Kenya. In Bangladesh, the interventions reduced antibiotic use most at the first measurement when children were on average 3 months old. Combining WSH and nutrition interventions did not reduce antibiotic use more than WSH or nutrition interventions alone. Our findings are broadly consistent with a recent double-blind, randomized controlled trial in urban Bangladesh that found that community-scale chlorination of drinking water reduced caregiver-reported antibiotic use by children in the past two months by 7%, along with a 23% reduction in diarrhea prevalence ^19^.

The observed reductions in antibiotic use demonstrate internal consistency with the trials’ previously reported intervention effects on diarrhea and respiratory illness. In Bangladesh, diarrhea was reduced by 31-38% in the WSH, nutrition and N+WSH arms compared to controls ^20^, while respiratory infections were reduced by 33% in the N+WSH arm but not the nutrition and WSH arms ^21^. Taken together, these findings support a causal explanation that WSH or nutrition interventions reduced pediatric antibiotic use by reducing illness that require antibiotic treatment. Among 14-month old children in the EED cohort in Bangladesh, TaqMan Array Card analysis of stool samples found that children in the WSH, nutrition and N+WSH groups carried fewer viruses and children in the WSH group carried fewer total pathogens than controls but there was no effect on carriage of bacterial pathogens in any intervention group ^24^. We found lower antibiotic use in all three of these study arms. Reductions in carriage of enteric viruses but not bacterial pathogens suggest that interventions may have reduced unnecessary antibiotic prescriptions prompted by viral infections.

In Kenya, the interventions had no effect on diarrhea ^22^, and respiratory infections were reduced by 13% in the nutrition arm during the first year of the trial but not during the full study period and not in the WSH and N+WSH arms ^23^. Similarly, in this analysis, total days of antibiotic use was reduced only in the nutrition arm, and only at the first measurement when children were on average 6 months old. However, lack of intervention effects on illness and antibiotic use in the N+WSH intervention group suggests that the observed effects in the nutrition group in Kenya should be interpreted with caution.

One possible explanation for different intervention effects in Bangladesh vs. Kenya is differential adherence. High intervention uptake was sustained throughout the study in Bangladesh ^25^, whereas in Kenya uptake was lower and further decreased later in the trial ^22^. In Bangladesh, in the WSH and N+WSH intervention groups, structured observations recorded defecation in hygienic latrines for 95-97% of adults, 85% of households had water and soap in latrine or kitchen areas, and 50-65% had detectable chlorine in their stored drinking water. In nutrition and N+WSH intervention groups, >80% of caregivers reported that children consumed the recommended amount of lipid-based nutrition supplements provided by the trial. In Kenya, 79-90% of WSH and N+WSH intervention households had improved latrines. Over 75% of households in these groups had water and soap at the handwashing location in the first year and approximately 20% in the second year of the trial. Approximately 40% of WSH and N+WSH intervention households had chlorine in their water in the first year and 20% in the second year of the trial. Adherence to consumption of recommended lipid-based nutrition supplements was >95% in the nutrition and N+WSH groups in both years of the trial. The observed reductions in antibiotic use in our analysis are consistent with these patterns in uptake and corroborate the importance of high intervention uptake in achieving effects on immediate as well as downstream outcomes.

Effects of WSH and nutrition interventions on community carriage of AMR need to be evaluated as an additional downstream outcome. Repeated antibiotic use within a short period persistently alters the gut microbiota into a predominantly resistant population ^26^. Children in LMICs often carry enteropathogens asymptomatically in their gut; when antibiotics are used to treat symptomatic respiratory and diarrheal infections, these “bystander” pathogens that are not the target of treatment are exposed to antibiotics, increasing the risk of resistance ^27^. We would expect the observed reductions in antibiotic use among children receiving interventions in our analysis to translate to reduced carriage of AMR. WASH interventions can additionally directly interrupt the environmental spread and transmission of antimicrobial resistant organisms ^28–30^.

Nadimpalli et al. (2022) found that community-scale water chlorination did not reduce antimicrobial resistance genes in child stool samples in urban Bangladesh despite reducing child diarrhea and reported antibiotic use, indicating that improving water quality alone in a setting with widespread contamination was not sufficient to reduce AMR ^31^. It has been suggested that poor sanitation ^3^ and environmental spread of antimicrobial resistant organisms ^32^ are stronger drivers of the global spread of AMR than antibiotic use. In a study of human gut metagenomes from 26 countries, access to improved drinking water and sanitation was associated with lower abundance of antimicrobial resistance genes ^33^. Diet is also associated with the occurrence of antimicrobial resistance genes in the gut ^34^, and nutrition improvements have been proposed to reduce AMR ^35^. Our findings support recommendations that future studies should assess the effect of WASH and nutrition interventions on community carriage of antimicrobial resistant organisms and antimicrobial resistance genes ^36^. These assessments can focus on sentinel organisms such as extended-spectrum beta-lactamase producing *E. coli*, following WHO recommendations for global AMR surveillance ^37^. Intervention studies should also collect data on antibiotic use as an outcome, which is cheaper and easier than measuring AMR in human biospecimens. There are no standardized data collection tools for recording antibiotic use. Studies can focus on predominantly used antibiotics in a given setting (e.g., based on pharmacy sales), and future work should explore best practices for developing and harmonizing data collection with respect to antibiotic classes of significance, optimal recall window for reporting use, and validation of self-reported use against medical records.

Additional potential downstream outcomes include effects on the microbiome and long-term sequelae. Diarrhea and antibiotic use are associated with alterations of gut microbiota and diminished microbiome richness and diversity ^38–40^. Children exposed to antibiotics are at increased risk of a range of conditions, including asthma, juvenile arthritis, type 1 diabetes, Crohn’s disease and mental illness; antibiotic-caused perturbations of the microbiome are believed to drive these risks ^40,41^. Interventions that reduce early life infections and antibiotic use can support the natural maturational development of microbiome composition from infancy through childhood and potentially offer protection against these sequelae. Notably, we observed the largest reductions in antibiotic use from the interventions among the youngest children (3-6 months) in our study. The gut microbiome experiences rapid changes between the ages of 3-14 months, undergoes transition between 15-31 months and starts to stabilize into a mostly adult-like composition between 31-36 months ^42^; minimizing perturbations to microbiota during these early life windows may deliver long-term health benefits.

A limitation of the study is that we relied on caregiver-reported antibiotic use, which is subject to poor recall and/or biased reporting given our non-blinded intervention. However, a previous study of birth cohorts in eight countries found good agreement between caregiver-reported antibiotic use in children and medical reports ^6^. Our findings are also internally consistent and biologically plausible; we observed reduced antibiotic use when high intervention uptake and reductions in diarrhea and respiratory infections were achieved. A strength of the study is evidence from two high-risk settings and cluster-randomized allocation of interventions with over 150 clusters in each trial’s substudy. The trials were efficacy trials with free provision of products and intensive promotion so the same effects may not be achieved when interventions are programmatically implemented at scale — nevertheless, they provide evidence that an effect on antibiotic use is possible with intensive WASH and nutritional interventions that reduce clinical illness among young children. We also note that the delivery mechanism for nutrition supplements was through home visits by project staff. Mass distribution of such supplements by healthcare systems can have different effects on antibiotic use. For example, adding nutrition supplements into health programs can increase program participation ^43,44^. While this can lead to higher immunization rates, which would reduce antibiotic use by reducing illness ^45^, increased contact with the health system can also increase unnecessary antibiotic prescriptions.

Global efforts to limit AMR primarily focus on limiting inappropriate antibiotic use, providing appropriate antimicrobials as needed, improving WASH access, and reducing exposure to untreated waste ^46,47^. While antibiotic stewardship can limit the unnecessary use of antibiotics and subsequent AMR ^48,49^, our results provide support for upstream WASH and nutrition interventions to reduce antibiotic use through decreased infection. Studies should assess the effects of water, sanitation, hygiene and nutrition interventions on community carriage of AMR.

## Methods

### Study design and participants

The WASH Benefits trials were cluster-randomized controlled trials that enrolled pregnant women in rural Bangladesh and Kenya. The Bangladesh trial (NCT01590095) was conducted in contiguous rural subdistricts in Gazipur, Mymensingh, Tangail and Kishoreganj districts of central Bangladesh. The Kenya trial (NCT01704105) was conducted in rural villages in Kakamega, Bungoma, and Vihiga counties in western Kenya. The study areas were chosen to have no other ongoing WASH/nutrition programs. Details of the study design have been described ^50^. Primary caregivers of children provided written informed consent. The study protocol was approved by human subjects committees at the International Centre for Diarrhoeal Disease Research, Bangladesh (PR11063), Kenya Medical Research Institute (SSC-2271), University of California, Berkeley (2011-09-3652), and Stanford University (25863).

### Randomization and masking

Field staff screened the areas for pregnant women. Geographically matched clusters of 6-8 compounds where the enrolled pregnant women lived were block-randomized to control or one of the intervention arms by an off-site investigator using a random number generator. The cluster design was chosen to minimize between-arm spillovers and facilitate intervention delivery logistics. Participants or data collectors were not blinded because interventions included visible hardware.

### Procedures

Interventions were initiated around when the birth cohort was born and included water treatment, sanitation, handwashing, nutrition, combined water treatment, sanitation and handwashing (WSH), and nutrition plus combined WSH (N+WSH). The water treatment component included chlorine tablets and a safe storage vessel provided to households in Bangladesh and chlorine dispensers installed at all community water locations plus bottled chlorine provided to households in Kenya. The sanitation component included double-pit latrines for all households in study compounds in Bangladesh, pit latrine upgrades with a reinforced slab and drop hole cover in Kenya, and child potties and hoes for feces management in both countries. The handwashing component included handwashing stations with soapy water solution and rinse water near the latrine and kitchen. The nutrition component included lipid-based nutrient supplements (LNS) for the birth cohort and age-appropriate recommendations on maternal nutrition and infant feeding practices.

Intervention hardware and consumables were provided free of charge and replenished throughout the study period. Local promoters hired and trained by study staff visited study compounds regularly to promote: (1) treating drinking water for children aged <36 months, (2) use of latrines/child potties for defecation and removal of human and animal feces from the compound, (3) handwashing with soap at critical times around food preparation, defecation, and contact with feces, and (4) LNS for children aged 6-24 months and age-appropriate nutrition practices from pregnancy to 24 months. In Bangladesh, promoters visited participants six times per month on average. In Kenya, they were instructed to visit participants several times in the first two months while interventions were delivered, monthly for the rest of the first year, and every two months the second year. In Bangladesh, promoters did not visit control households (passive controls). In Kenya, they visited control households (active controls) monthly to measure child mid-upper arm circumference. User uptake of targeted behaviors was assessed by structured and spot check observations. Uptake was high and sustained in Bangladesh ^25,51,52^ but lower and variable in Kenya ^22^. Details of intervention delivery and uptake have been previously described ^22,25,51,52^.

Antibiotic use was recorded among children participating in a substudy conducted to assess environmental enteric dysfunction (EED). The EED substudy was conducted in the control, nutrition, WSH and N+WSH arms of the parent trial. To facilitate collection of biological specimens, enrollment in the substudy focused on areas close to the field laboratory and did not follow the geographic matching of the parent trial. The EED substudy in Kenya was limited to the Kakamega and Bungoma counties and excluded children <1 month old without a clinic card and children <2 weeks old due to lack of parental consent. The EED substudy enrolled 267 clusters in Bangladesh and 190 clusters in Kenya and aimed to enroll 1500 children (375 per arm) from the birth cohort per country.

For children in the EED substudy, we recorded antibiotic use in three longitudinal rounds in approximately one-year intervals. Due to challenging logistics and political unrest, we were not able to synchronize child ages at follow-up across the countries; children were on average 3, 14, and 28 months old in Bangladesh and 6, 17, and 22 months old in Kenya at the three measurement points. Trained field staff asked the primary caregiver how many times the child used antibiotics within 90 days before the visit, and the number of days the child used specific antibiotics during this window. When available, field staff asked for the prescription or the packaging for the antibiotic, and otherwise asked caregivers to recall the name of the antibiotic from among a list of 11 antibiotics commonly used in the study regions (cotrimoxazole, amoxicillin, flucloxacillin, ciprofloxacin, erythromycin, azithromycin, nalidixic acid, doxycycline, penicillin, chloramphenicol, metronidazole). If the taken antibiotic was not listed, field staff asked caregivers to specify the antibiotic or else marked it as unknown.

### Outcomes

For the present analysis, our pre-specified primary outcome was the prevalence of children who used antibiotics at least once within 90 days prior to data collection. Pre-specified secondary outcomes were the prevalence of children who used antibiotics multiple times, number of episodes of antibiotic use, and number of days of antibiotic use, within 90 days prior to data collection. We compared these outcomes in each intervention arm against controls and in the N+WSH arm against the nutrition and WSH arms.

### Statistical analysis

*Hypotheses*. We hypothesized that (1) children receiving nutrition, WSH or N+WSH interventions would have reduced antibiotic use compared to controls and (2) children receiving N+WSH interventions would have reduced antibiotic use compared to those receiving WSH or nutrition interventions alone. These hypotheses follow the pre-specified hypotheses of the WASH Benefits trials for the primary trial outcomes.

*Estimation strategy*. We conducted comparisons separately for each country, using pooled data from all three measurement points. Analyses were intention-to-treat. We estimated prevalence ratios (PRs) and prevalence differences (PDs) for binary outcomes, and count ratios (CRs) and count differences (CDs) for count outcomes. We used generalized linear models with robust standard errors to account for geographical clustering and repeated measurements. We used a Poisson error distribution and log link to estimate PRs for binary outcomes ^53^, Poisson or negative binomial error distribution and log link to estimate CRs for count outcomes, and a Gaussian error distribution and identity link to estimate PDs and CDs. Randomization led to good balance in measured covariates across arms ^20,22^ so our primary inference focused on unadjusted comparisons between groups. In additional analyses, we estimated adjusted effects by including variables strongly associated with the outcome to potentially improve the precision of our estimates ^54^. As per the analysis plan of the WASH Benefits trial ^55^ (updated on 2016.02.05, https://osf.io/63mna/), we considered the following adjustment covariates: data collection date (in 3-month intervals), child age, sex and birth order, mother’s age, height and education, household food insecurity, number of individuals <18 years in household, number of individuals living in compound, distance to household’s primary drinking water source, housing materials, and household wealth index calculated from principal components analysis of household assets. We used likelihood ratio tests to assess the association between each covariate and outcome and included covariates with a p-value<0.20 in adjusted analyses. Details of our analysis approach are available in a pre-specified analysis plan, along with de-identified datasets and analysis scripts (https://osf.io/t7fmw/). Analyses were conducted using R software (version 4.0.3 GUI 1.73). We followed CONSORT reporting guidelines (Table S1).

*Statistical power.* We calculated the minimum detectable effects (MDEs) based on the prevalence of control children who used antibiotics at least once in the 90 days (63% in Bangladesh, 53% in Kenya), number of observations per study cluster (17 in Bangladesh, 26 in Kenya), and the intracluster correlation coefficient for observations in the same cluster (0.04 in Bangladesh, 0.03 in Kenya). Our sample size yields 80% power with a two-sided alpha of 0.05 to detect the following MDEs between any intervention arm vs. controls: 11% relative reduction in Bangladesh and 13% relative reduction in Kenya in the prevalence of using antibiotics at least once in 90 days.

*Effect modification.* Children of different age groups have different physiological characteristics, levels of immunity and risk of infection ^56^. In addition, different stages of breastfeeding, weaning, mobility and dexterity result in varying exposure to pathogens through contaminated water, food, hands and objects ^57–59^. Child sex may influence intervention delivery, immune status and antibiotic use, due to biological differences, differential treatment by caregivers, or sex-specific behaviors ^60–62^. We hypothesized that the effects of WSH and nutrition interventions on antibiotic use could vary with child age and sex. Within each country, we investigated effect modification by the three measurement points (corresponding to a mean child age of 3, 14, 28 months in Bangladesh, and 6, 17, 22 months in Kenya) and by sex. We included interaction terms between study arm and these effect modifiers in regression models and investigated both multiplicative and additive interaction. We interpreted interaction p-values<0.20 as evidence of effect modification.

*Sensitivity analyses*. It is possible that caregiver-reported antibiotic use within the last three months is subject to inaccurate recall, and a shorter recall window may increase the accuracy of reporting. As sensitivity analyses, we estimated intervention effects on antibiotic use during the last month and last two weeks. These variables were derived from a separate question on how long ago the child last used antibiotics. In Kenya, this question recorded both antibiotics and non-antibiotic medications so the sensitivity analysis was only conducted for Bangladesh.

## Data Availability

Upon publication, all data will be available on Open Science Framework at https://osf.io/t7fmw/

## Acknowledgments

This research was financially supported by a global development grant (OPPGD759) from the Bill & Melinda Gates Foundation to the University of California, Berkeley, CA, USA.

## Contributors

AE: Conceptualisation, data curation, formal analysis, methodology, writing – original draft

ANM: Data curation, formal analysis, writing – review & editing

ZBD: Visualisation, writing – review & editing

DKJ: Investigation, writing – review & editing

SA: Supervision, data curation, investigation, writing – review & editing

BSA: Supervision, data curation, investigation, writing – review & editing

GR: Supervision, data curation, investigation, writing – review & editing

CH: Investigation, writing – review & editing

AJP: Investigation, supervision, writing – review & editing

CPS: Investigation, supervision, writing – review & editing

STT: Data curation, investigation, writing – review & editing

JAG: Data curation, investigation, writing – review & editing

JBC: Investigation, project administration, writing – review & editing

MW: Data curation, investigation, writing – review & editing

GH: Investigation, writing – review & editing

MZR: Data curation, investigation, writing – review & editing

CDA: Data curation, investigation, writing – review & editing

HND: Supervision, investigation, writing – review & editing

SMN: Investigation, supervision, writing – review & editing

DM: Supervision, investigation, writing – review & editing

BC: Data curation, investigation, writing – review & editing

MN: Writing – review & editing

MAI: Writing – review & editing

AEH: Methodology, writing – review & editing

CN: Investigation, supervision, writing – review & editing

LU: Investigation, supervision, writing – review & editing

MR: Investigation, supervision, writing – review & editing

JMC: Conceptualisation, funding acquisition, investigation, supervision, writing – review & editing

SPL: Conceptualisation, funding acquisition, investigation, supervision, writing – review & editing

BFA: Conceptualisation, funding acquisition, investigation, methodology, writing – review & editing

AL: Conceptualisation, supervision, data curation, investigation, writing – review & editing

