## Supplemental Information for "Can drinking water, sanitation, handwashing, and nutritional interventions reduce antibiotic use in young children?"

### Table of Contents


| Item | Pages |
|---|---|
| **Table S1.** CONSORT Checklist | 2-3 |
| **Table S2.** Enrollment characteristics by intervention group within the EED substudy and the WASH Benefits parent trial in Bangladesh | 4 |
| **Table S3**. Enrollment characteristics by intervention group within the EED substudy and the WASH Benefits parent trial in Kenya | 5 |
| **Table S4.** Enrollment characteristics of children in the EED substudy included in follow-up vs. lost to follow-up in Bangladesh | 6 |
| **Table S5.** Enrollment characteristics of children in the EED substudy included in follow-up vs. lost to follow-up in Kenya | 7 |
| **Table S6.** Caregiver-reported antibiotic use in last 3 months by young children in subgroups of age and sex, Bangladesh | 8 |
| **Table S7.** Caregiver-reported antibiotic use in last 3 months by young children in subgroups of age and sex, Kenya | 9 |
| **Table S8.** WSH, nutrition and N+WSH interventions vs. controls on antibiotic use within last 3 months among children in Bangladesh | 10 |
| **Table S9.** Combined N+WSH intervention vs. WSH and nutrition interventions on antibiotic use within last 3 months among children in Bangladesh | 11 |
| **Table S10.** WSH, nutrition and N+WSH interventions vs. controls on antibiotic use within last 3 months among children in Kenya | 12 |
| **Table S11.** Combined N+WSH intervention vs. WSH and nutrition interventions on antibiotic use within last 3 months among children in Kenya | 13 |
| **Table S12.** Multiplicative interaction by age and sex for unadjusted intervention effects on antibiotic use within last 3 months, Bangladesh | 14 |
| **Table S13.** Additive interaction by age and sex for unadjusted intervention effects on antibiotic use within last 3 months, Bangladesh | 15 |
| **Table S14.** Multiplicative interaction by age and sex for unadjusted intervention effects on antibiotic use within last 3 months, Kenya | 16 |
| **Table S15.** Additive interaction by age and sex for unadjusted intervention effects on antibiotic use within last 3 months, Kenya | 17 |
| **Table S16.** Sensitivity analysis, unadjusted intervention effects on antibiotic use within last 2 weeks and last month, Bangladesh* | 18 |
| **Figure S1.** CONSORT Diagram for the WASH Benefits Bangladesh environmental enteric dysfunction substudy | 19 |
| **Figure S2.** CONSORT Diagram for the WASH Benefits Kenya environmental enteric dysfunction substudy | 20 |

**Table S1.** CONSORT Checklist

| Item | Description | Reported in Section |
|---|---|---|
| **Title and Abstract** | | |
| 1a | Identification as a randomized trial in the title; Identification as a cluster randomized trial in the title | Title |
| 1b | Structured summary of trial design, methods, results, and conclusions | Abstract |
| **Introduction** | | |
| Background and Objectives | | |
| 2a | Scientific background and explanation of rationale; Rationale for using a cluster design | Introduction (paragraph 1) |
| 2b | Specific objectives or hypotheses; Whether objectives pertain to the cluster level, the individual participant level, or both | Introduction (paragraph 2) |
| **Methods** | | |
| Trial Design | | |
| 3a | Description of trial design (such as parallel, factorial) including allocation ratio; Definition of cluster and description of how the design features apply to the clusters | Methods: Study design, Data collection (paragraph 1) |
| 3b | Important changes to methods after trial commencement (such as eligibility criteria), with reasons | N/A |
| Participants | | |
| 4a | Eligibility criteria for participants; Eligibility criteria for clusters | Methods: Study design |
| 4b | Settings and locations where the data were collected | Methods: Study design |
| Interventions | | |
| 5 | The interventions for each group with sufficient details to allow replication, including how and when they were actually administered; Whether interventions pertain to the cluster level, the individual participant level, or both | Methods: Study design, Interventions (paragraph 1) |
| Outcomes | | |
| 6a | Completely defined pre-specified primary and secondary outcome measures, including how and when they were assessed; Whether outcome measures pertain to the cluster level, the individual participant level, or both | Methods: Data collection (paragraph 2), Statistical parameters and estimation strategy (paragraph 1) |
| 6b | Any changes to trial outcomes after the trial commenced, with reasons | N/A |
| Sample Size | | |
| 7a | How sample size was determined; Method of calculation, number of cluster(s) (and whether equal or unequal cluster sizes are assumed), cluster size, a coefficient of intracluster correlation (ICC or k), and an indication of its uncertainty | Methods: Minimum detectable effect size |
| 7b | When applicable, explanation of any interim analyses and stopping guidelines | N/A |
| **Randomization** | | |
| Sequence Generation | | |
| 8a | Method used to generate the random allocation sequence | Methods: Study design |
| 8b | Type of randomization; details of any restriction (such as blocking and block size); Details of stratification or matching if used | Methods: Study design |
| Allocation Concealment Mechanism | | |
| 9 | Mechanism used to implement the random allocation sequence (such as sequentially numbered containers), describing any steps taken to conceal the sequence until interventions were assigned; Specification that allocation was based on clusters rather than individuals and whether allocation concealment (if any) was at the cluster level, the individual participant level, or both | Methods: Study design |
| Implementation | | |
| 10a | Who generated the random allocation sequence, who enrolled clusters, and who assigned clusters to interventions | Methods: Study design |
| 10b | Mechanism by which individual participants were included in clusters for the purposes of the trial (such as complete enumeration, random sampling) | Methods: Study design |
| 10c | From whom consent was sought (representatives of the cluster, or individual cluster members, or both) and whether consent was sought before or after randomization | Methods: Ethics |
| Blinding | | |
| 11a | If done, who was blinded after assignment to interventions (for example, participants, care providers, those assessing outcomes) and how | Methods: Data collection (paragraph 2) |
| 11b | If relevant, description of the similarity of interventions | N/A |

| Item | Description | Reported in Section |
|---|---|---|
| Statistical Methods | | |
| 12a | Statistical methods used to compare groups for primary and secondary outcomes; How clustering was taken into account | Methods: Statistical parameters and estimation strategy (paragraph 2) |
| 12b | Methods for additional analyses, such as subgroup analyses and adjusted analyses | Methods: Statistical parameters and estimation strategy (paragraph 2), Subgroup analyses, Sensitivity analyses |
| **Results** | | |
| Participant Flow | | |
| 13a | For each group, the numbers of participants/clusters who were randomly assigned, received intended treatment, and were analyzed for the primary outcome | Results: Enrollment, Fig S1, Fig S2 |
| 13b | For each group, losses and exclusions after randomization, together with reasons, for both clusters and individual cluster members | Results: Enrollment, Fig S1, Fig S2 |
| Recruitment | | |
| 14a | Dates defining the periods of recruitment and follow-up | Results: Enrollment |
| 14b | Why the trial ended or was stopped | N/A |
| Baseline Data | | |
| 15 | A table showing baseline demographic and clinical characteristics for each group; Baseline characteristics for the individual and cluster levels as applicable for each group | Tables S2, S3 |
| Numbers Analyzed | | |
| 16 | For each group, number of participants/clusters (denominator) included in each analysis and whether the analysis was by the original assigned groups | Statistical parameters and estimation strategy (paragraph 2), Tables S6, S7 |
| Outcomes and Estimation | | |
| 17a | For each primary and secondary outcome, results for each group, and the estimated effect size and its precision (such as 95% confidence interval); Results at the individual and cluster levels as applicable and a coefficient of intracluster correlation (ICC or k) for each primary outcome | Results: Intervention effects on antibiotic use, Fig 1, Fig2, Tables S8-S11 |
| 17b | For binary outcome, presentation of both absolute and relative effect sizes is recommended | Results: Intervention effects on antibiotic use, Tables S8-S11 |
| Ancillary Analyses | | |
| 18 | Results of any other analyses performed, including subgroup analyses and adjusted analyses, distinguishing pre-specified from exploratory | Results: Subgroup analyses, Secondary and sensitivity analyses, Fig 3, Fig 4, Tables S12-S16 |
| Harms | | |
| 19 | All important harms or unintended effects in each group | N/A |
| **Discussion** | | |
| Limitations | | |
| 20 | Trial limitations, addressing sources of potential bias, imprecision and, if relevant, multiplicity of analyses | Discussion |
| Generalizability | | |
| 21 | Generalizability (external validity, applicability) of the trial findings; Generalizability to clusters and/or individual participants (as relevant) | Discussion |
| Interpretation | | |
| 22 | Interpretation consistent with results, balancing benefits and harms, and considering other relevant evidence | Discussion |
| **Other Information** | | |
| Registration | | |
| 23 | Registration number and name of trial registry | Methods: Study design |
| Protocol | | |
| 24 | Where the full trial protocol can be accessed, if available | N/A |
| Funding | | |
| 25 | Sources of funding and other support (such as supply of drugs), role of funders | Funding |

**Table S2.** Enrollment characteristics by intervention group within the EED substudy and the WASH Benefits parent trial in Bangladesh

| | EED substudy | | | | WASH Benefits parent trial | | | |
|---|---|---|---|---|---|---|---|---|
| | C | WSH | N | N+WSH | C | WSH | N | N+WSH |
| Number of compounds | 454 | 429 | 450 | 450 | 1382 | 698 | 699 | 686 |
| **Socio-demographics** | | | | | | | | |
| Mother's age in years, mean (SD) | 23 (5) | 24 (5) | 24 (5) | 24 (5) | 24 (5) | 24 (5) | 24 (5) | 24 (6) |
| Mother's years of education, mean (SD) | 7 (3) | 6 (3) | 6 (4) | 6 (3) | 6 (3) | 6 (3) | 6 (3) | 6 (3) |
| Father's years of education, mean (SD) | 5 (4) | 5 (4) | 5 (4) | 5 (4) | 5 (4) | 5 (4) | 5 (4) | 5 (4) |
| Father works in agriculture, % (n) | 23% (104) | 29% (128) | 34% (148) | 28% (127) | 30% (414) | 31% (216) | 33% (232) | 30% (207) |
| Number of people in household, mean (SD) | 5 (2) | 5 (2) | 5 (2) | 5 (2) | 5 (2) | 5 (2) | 5 (2) | 5 (2) |
| Has electricity, % (n) | 60% (269) | 62% (278) | 62% (269) | 61% (272) | 57% (784) | 61% (426) | 59% (409) | 60% (412) |
| Has cement floor, % (n) | 17% (75) | 12% (55) | 11% (50) | 12% (53) | 10% (145) | 11% (77) | 10% (67) | 10% (72) |
| Acres of agricultural land owned, mean (SD) | 0 (0) | 0 (0) | 0 (0) | 0 (0) | 0.15 (0.21) | 0.15 (0.23) | 0.16 (0.27) | 0.14 (0.38) |
| **Drinking water** | | | | | | | | |
| Primary water source is shallow tubewell, % (n) | 73% (329) | 76% (338) | 71% (309) | 71% (318) | 75% (1038) | 78% (546) | 74% (519) | 73% (504) |
| Stored water observed at home, % (n) | 51% (230) | 45% (200) | 48% (209) | 51% (229) | 48% (666) | 43% (304) | 43% (301) | 48% (331) |
| Reported treating water yesterday, % (n) | 0% (1) | 0% (0) | 0% (0) | 0% (1) | 0% (4) | 0% (0) | 0% (0) | 0% (2) |
| Distance to water source in minutes, mean (SD) | 1 (2) | 1 (6) | 1 (2) | 1 (2) | 1 (3) | 1 (5) | 1 (3) | 1 (2) |
| **Sanitation** | | | | | | | | |
| Reported daily open defecation | | | | | | | | |
| Adult men, % (n) | 4% (19) | 7% (30) | 9% (38) | 9% (38) | 7% (97) | 8% (54) | 9% (59) | 7% (50) |
| Adult women, % (n) | 3% (12) | 4% (16) | 5% (23) | 5% (21) | 4% (62) | 4% (29) | 6% (39) | 4% (24) |
| Children 8-15 yrs, % (n) | 5% (9) | 8% (17) | 8% (13) | 11% (22) | 10% (53) | 10% (30) | 8% (23) | 10% (28) |
| Children 3-8 yrs, % (n) | 30% (64) | 37% (90) | 40% (90) | 37% (92) | 38% (267) | 38% (137) | 39% (129) | 37% (134) |
| Children <3 yrs, % (n) | 72% (71) | 75% (73) | 80% (68) | 88% (79) | 82% (245) | 79% (123) | 85% (128) | 88% (123) |
| Latrine | | | | | | | | |
| Owned by household, % (n) | 60% (271) | 55% (244) | 54% (234) | 51% (230) | 54% (750) | 53% (373) | 54% (377) | 53% (367) |
| Has concrete slab, % (n) | 97% (426) | 93% (401 | 93% (382) | 94% (399) | 95% (1251) | 93% (620) | 94% (620) | 94% (621) |
| Has functional water seal, % (n) | 38% (157) | 26% (95) | 32% (114) | 31% (111) | 31% (358) | 26% (152) | 31% (183) | 27% (155) |
| Owns child potty | 8% (37) | 4% (20) | 6% (27) | 5% (21) | 4% (61) | 4% (27) | 5% (36) | 4% (30) |
| Human feces observed | | | | | | | | |
| In house, % (n) | 6% (25) | 8% (36) | 9% (41) | 8% (36) | 8% (114) | 7% (48) | 8% (58) | 7% (49) |
| In child's play area, % (n) | 1% (5) | 1% (4) | 2% (7) | 1% (6) | 2% (21) | 1% (7) | 1% (8) | 1% (7) |
| **Handwashing** | | | | | | | | |
| Within 6 steps of latrine | | | | | | | | |
| Has water, % (n) | 21% (84) | 13% (51) | 10% (38) | 13% (54) | 14% (178) | 10% (67) | 10% (62) | 11% (72) |
| Has soap, % (n) | 7% (88) | 7% (42) | 5% (32) | 6% (36) | 11% (45) | 8% (32) | 6% (23) | 7% (27) |
| Within 6 steps of kitchen | | | | | | | | |
| Has water, % (n) | 12% (48) | 10% (4) | 11% (43) | 10% (42) | 9% (118) | 9% (61) | 9% (61) | 9% (60) |
| Has soap, % (n) | 4% (18) | 3% (11) | 5% (19) | 3% (14) | 3% (33) | 2% (15) | 4% (23) | 3% (18) |
| **Nutrition** | | | | | | | | |
| Household is food secure, % (n) | 71% (485) | 74% (331) | 67% (298) | 71% (308) | 67% (932) | 69% (482) | 69% (479) | 71% (485) |

**Table S3**. Enrollment characteristics by intervention group within the EED substudy and the WASH Benefits parent trial in Kenya

| | EED substudy | | | | WASH Benefits parent trial | | | |
|---|---|---|---|---|---|---|---|---|
| | C | WSH | N | N+WSH | C | WSH | N | N+WSH |
| Number of compounds | 511 | 456 | 434 | 448 | 1919 | 912 | 843 | 921 |
| **Socio-demographics** | | | | | | | | |
| Mother's age in years, mean (SD) | 26 (6) | 26 (6) | 26 (6) | 26 (6) | 26 (6) | 26 (6) | 26 (6) | 26 (6) |
| Mother completed at least primary education, % (n) | 45% (227) | 44% (200) | 50% (215) | 49% (217) | 48% (916) | 47% (430) | 49% (409) | 48% (438) |
| Father completed at least primary education, % (n) | 59% (279) | 59% (252) | 61% (248) | 62% (258) | 62% (1098) | 61% (521) | 64% (491) | 62% (526) |
| Father works in agriculture, % (n) | 47% (231) | 45% (197) | 49% (201) | 44% (187) | 41% (749) | 43% (374) | 43% (343) | 43% (372) |
| Number of people in compound, mean (SD) | 9 (6) | 9 (5) | 9 (6) | 9 (6) | 8 (5) | 8 (5) | 8 (7) | 8 (5) |
| Has electricity, % (n | 4% (22) | 8% (35) | 7% (30) | 6% (25) | 6% (122) | 7% (64) | 7% (58) | 7% (67) |
| Has cement floor, % (n) | 5% (25) | 5% (25) | 5% (20) | 4% (19) | 6% (107) | 5% (50) | 6% (48) | 6% (56) |
| Has iron roof, % (n) | 60% (305) | 59% (271) | 66% (285) | 62% (279) | 68% (1302) | 63% (574) | 69% (581) | 67% (615) |
| **Drinking water** | | | | | | | | |
| Primary water source is improved, % (n) | 79% (402) | 71% (324) | 71% (308) | 80% (355) | 76% (1446) | 69% (624) | 72% (603) | 76% (697) |
| Reported treating currently stored water, % (n) | 10% (43) | 13% (46) | 8% (27) | 13% (46) | 13% (196) | 13% (97) | 12% (79) | 14% (106) |
| Distance to water source in minutes, mean (SD) | 10 (10) | 10 (11) | 10 (11) | 10 (12) | 11 (12) | 11 (13) | 11 (12) | 11 (12) |
| **Sanitation** | | | | | | | | |
| Children <3 yrs daily defecate in the open, % (n) | 77% (230) | 76% (195) | 78% (188) | 79% (187) | 78% (789) | 77% (394) | 79% (363) | 78% (388) |
| Household owns latrine, % (n) | 81% (415) | 86% (390) | 85% (370) | 86% (384) | 82% (1561) | 83% (754) | 83% (701) | 83% (764) |
| Access to improved latrine, % (n) | 17% (82) | 17% (75) | 16% (66) | 15% (65) | 17% (309) | 18% (153) | 15% (119) | 16% (143) |
| Human feces observed in compound, % (n) | 7% (36) | 9% (39) | 9% (40) | 9% (42) | 9% (163) | 8% (73) | 9% (73) | 9% (87) |
| **Handwashing location** | | | | | | | | |
| Has water within 2 m, % (n) | 24% (121) | 28% (127) | 27% (117) | 27% (121) | 25% (487) | 28% (251) | 27% (228) | 27% (249) |
| Has soap within 2 m, % (n) | 8% (40) | 13% (61) | 11% (46) | 10% (46) | 9% (164) | 13% (115) | 11% (90) | 9% (87) |
| **Nutrition** | | | | | | | | |
| Moderate to severe household hunger, % (n) | 11% (55) | 9% (41) | 12% (54) | 11% (47) | 11% (203) | 11% (101) | 12% (98) | 11% (104) |

**Table S4.** Enrollment characteristics of children in the EED substudy included in follow-up vs. lost to follow-up in Bangladesh

| | Included | Missing from follow-up at age 14 months | Missing from follow-up at age 28 months |
|---|---|---|---|
| Number of compounds | 1778 | 284 | 273 |
| **Socio-demographics** | | | |
| Mother's age in years, mean (SD) | 24 (5) | 24 (5) | 24 (5) |
| Mother's years of education, mean (SD) | 6 (3) | 6 (3) | 6 (3) |
| Father's years of education, mean (SD) | 5 (4) | 5 (4) | 5 (4) |
| Father works in agriculture, % (n) | 29% (507) | 24% (69) | 26% (70) |
| Number of people in household, mean (SD) | 5 (2) | 5 (2) | 5 (2) |
| Has electricity, % (n) | 61% (1088) | 60% (169) | 56% (153) |
| Has cement floor, % (n) | 13% (233) | 14% (40) | 15% (42) |
| Acres of agricultural land owned, mean (SD) | 0 (0) | 0 (0) | 0 (0) |
| **Drinking water** | | | |
| Primary water source is shallow tubewell, % (n) | 73% (1294) | 70% (200) | 70% (191) |
| Stored water observed at home, % (n) | 49% (868) | 51% (144) | 52% (142) |
| Reported treating water yesterday, % (n) | 0% (2) | 0% (0) | 0% (0) |
| Distance to water source in minutes, mean (SD) | 1 (3) | 1 (2) | 1 (1) |
| **Sanitation** | | | |
| Reported daily open defecation | | | |
| Adult men, % (n) | 7% (125) | 8% (21) | 7% (18) |
| Adult women, % (n) | 4% (72) | 5% (14) | 3% (9) |
| Children 8-15 yrs, % (n) | 8% (61) | 11% (12) | 14% (15) |
| Children 3-8 yrs, % (n) | 36% (337) | 39% (57) | 37% (51) |
| Children <3 yrs, % (n) | 79% (291) | 83% (45) | 81% (38) |
| Latrine | | | |
| Owned by household, % (n) | 55% (979) | 52% (148) | 56% (154) |
| Has concrete slab, % (n) | 94% (1608) | 96% (258) | 95% (249) |
| Has functional water seal, % (n) | 32% (477) | 31% (73) | 36% (84) |
| Owns child potty | 6% (105) | 7% (20) | 7% (20) |
| Human feces observed | | | |
| In house, % (n) | 8% (138) | 4% (12) | 7% (18) |
| In child's play area, % (n) | 1% (22) | 1% (3) | 2% (6) |
| **Handwashing** | | | |
| Within 6 steps of latrine | | | |
| Has water, % (n) | 14% (227) | 16% (42) | 16% (40) |
| Has soap, % (n) | 8% (127) | 10% (27) | 11% (27) |
| Within 6 steps of kitchen | | | |
| Has water, % (n) | 11% (173) | 12% (30) | 9% (21) |
| Has soap, % (n) | 4% (62) | 5% (12) | 2% (5) |
| **Nutrition** | | | |
| Household is food secure, % (n) | 71% (1254) | 68% (192) | 68% (187) |

**Table S5.** Enrollment characteristics of children in the EED substudy included in follow-up vs. lost to follow-up in Kenya

| | Included | Missing from follow-up at age 17 months | Missing from follow-up at age 22 months |
|---|---|---|---|
| Number of compounds | 1849 | 462 | 556 |
| **Socio-demographics** | | | |
| Mother's age in years, mean (SD) | 26 (6) | 25 (6) | 25 (6) |
| Mother completed at least primary education, % (n) | 47% (859) | 49% (225) | 49% (271) |
| Father completed at least primary education, % (n) | 60% (1037) | 62% (267) | 61% (310) |
| Father works in agriculture, % (n) | 46% (816) | 42% (181) | 43% (223) |
| Number of people in compound, mean (SD) | 9 (6) | 8 (6) | 8 (5) |
| Has electricity, % (n) | 6% (112) | 7% (31) | 6% (33) |
| Has cement floor, % (n) | 5% (89) | 6% (27) | 5% (29) |
| Has iron roof, % (n) | 62% (1140) | 60% (279) | 61% (341) |
| **Drinking water** | | | |
| Primary water source is improved, % (n) | 75% (1389) | 74% (340) | 76% (424) |
| Reported treating currently stored water, % (n) | 11% (162) | 11% (39) | 12% (54) |
| Distance to water source in minutes, mean (SD) | 10 (11) | 10 (9) | 10 (11) |
| **Sanitation** | | | |
| Children <3 yrs daily defecate in the open, % (n) | 78% (800) | 77% (178) | 71% (205) |
| Household owns latrine, % (n) | 84% (1559) | 81% (375) | 82% (454) |
| Access to improved latrine, % (n) | 16% (288) | 19% (83) | 18% (93) |
| Human feces observed in compound, % (n) | 9% (157) | 9% (40) | 10% (55) |
| **Handwashing location** | | | |
| Has water within 2 m, % (n) | 26% (486) | 26% (122) | 27% (148) |
| Has soap within 2 m, % (n) | 10% (193) | 11% (52) | 10% (58) |
| **Nutrition** | | | |
| Moderate to severe household hunger, % (n) | 11% (197) | 14% (65) | 11% (59) |

**Table S6.** Caregiver-reported antibiotic use in last 3 months by young children in subgroups of age and sex, Bangladesh

| | All observations | | Mean child age | | | | | | Sex | | | |
|---|---|---|---|---|---|---|---|---|---|---|---|---|
| | | | 3 months | | 14 months | | 28 months | | Female | | Male | |
| Arm | N | % (n) | N | % (n) | N | % (n) | N | % (n) | N | % (n) | N | % (n) |
| Used antibiotics ≥1 time | | | | | | | | | | | | |
| C | 1005 | 63.2 (635) | 270 | 54.1 (146) | 377 | 75.1 (283) | 358 | 57.5 (206) | 515 | 62.3 (321) | 490 | 64.1 (314) |
| WSH | 1077 | 57.0 (614) | 285 | 45.6 (130) | 395 | 70.6 (279) | 397 | 51.6 (205) | 510 | 54.7 (279) | 567 | 59.1 (335) |
| N | 1018 | 54.1 (551) | 269 | 41.3 (111) | 377 | 68.2 (257) | 372 | 49.2 (183) | 492 | 49.6 (244) | 526 | 58.4 (307) |
| N+WSH | 1058 | 54.2 (573) | 278 | 36.0 (100) | 379 | 71.5 (271) | 401 | 50.4 (202) | 565 | 49.7 (281) | 493 | 59.2 (292) |
| Used antibiotics ≥2 times | | | | | | | | | | | | |
| C | 1005 | 24.7 (248) | 270 | 24.8 (67) | 377 | 34.8 (131) | 358 | 14.0 (50) | 515 | 22.1 (114) | 490 | 27.4 (134) |
| WSH | 1077 | 18.2 (196) | 285 | 14.0 (40) | 395 | 25.6 (101) | 397 | 13.9 (55) | 510 | 15.9 (81) | 567 | 20.3 (115) |
| N | 1018 | 16.4 (167) | 269 | 12.6 (34) | 377 | 24.4 (92) | 372 | 11.0 (41) | 492 | 15.7 (77) | 526 | 17.1 (90) |
| N+WSH | 1058 | 16.2 (171) | 278 | 11.9 (33) | 379 | 26.9 (102) | 401 | 9.0 (36) | 565 | 14.5 (82) | 493 | 18.1 (89) |
| Arm | N | Mean (SD) | N | Mean (SD) | N | Mean (SD) | N | Mean (SD) | N | Mean (SD) | N | Mean (SD) |
| Episodes of antibiotic use | | | | | | | | | | | | |
| C | 1005 | 0.98 (0.98) | 270 | 0.90 (1.06) | 377 | 1.23 (0.99) | 358 | 0.77 (0.84) | 515 | 0.92 (0.94) | 490 | 1.03 (1.02) |
| WSH | 1077 | 0.81 (0.88) | 285 | 0.65 (0.87) | 395 | 1.05 (0.93) | 397 | 0.68 (0.78) | 510 | 0.74 (0.81) | 567 | 0.87 (0.93) |
| N | 1018 | 0.77 (0.90) | 269 | 0.58 (0.82) | 377 | 1.04 (0.99) | 372 | 0.64 (0.79) | 492 | 0.70 (0.86) | 526 | 0.84 (0.93) |
| N+WSH | 1058 | 0.77 (0.89) | 278 | 0.56 (0.93) | 379 | 1.08 (0.95) | 401 | 0.61 (0.71) | 565 | 0.70 (0.87) | 493 | 0.84 (0.91) |
| Total days of antibiotic use | | | | | | | | | | | | |
| C | 995 | 5.06 (5.81) | 265 | 4.77 (6.36) | 373 | 6.27 (5.91) | 357 | 4.01 (4.99) | 511 | 4.77 (5.58) | 484 | 5.37 (6.03) |
| WSH | 1075 | 4.12 (5.05) | 283 | 3.54 (5.18) | 395 | 5.13 (5.20) | 397 | 3.52 (4.64) | 509 | 3.82 (4.83) | 566 | 4.39 (5.22) |
| N | 1016 | 4.02 (5.30) | 268 | 3.00 (4.84) | 376 | 5.34 (5.74) | 372 | 3.41 (4.86) | 492 | 3.67 (5.15) | 524 | 4.34 (5.41) |
| N+WSH | 1055 | 4.05 (5.39) | 276 | 3.25 (6.01) | 379 | 5.70 (5.88) | 400 | 3.04 (3.86) | 564 | 3.65 (5.14) | 491 | 4.52 (5.63) |

C: Control; WSH: Water, sanitation, hygiene; N: Nutrition, N+WSH: Nutrition+ water, sanitation, hygiene; SD: Standard deviation.

**Table S7.** Caregiver-reported antibiotic use in last 3 months by young children in subgroups of age and sex, Kenya

| | All observations | | Mean child age | | | | | | Sex | | | |
|---|---|---|---|---|---|---|---|---|---|---|---|---|
| | | | 6 months | | 17 months | | 22 months | | Female | | Male | |
| Arm | N | % (n) | N | % (n) | N | % (n) | N | % (n) | N | % (n) | N | % (n) |
| Used antibiotics ≥1 time | | | | | | | | | | | | |
| C | 1143 | 52.6 (601) | 360 | 52.5 (189) | 394 | 53.3 (210) | 389 | 51.9 (202) | 571 | 53.2 (304) | 572 | 51.9 (297) |
| WSH | 1043 | 50.3 (525) | 366 | 49.7 (182) | 345 | 53.9 (186) | 332 | 47.3 (157) | 568 | 49.5 (281) | 475 | 51.4 (244) |
| N | 1048 | 50.4 (528) | 347 | 49.3 (171) | 358 | 54.8 (196) | 343 | 46.9 (161) | 495 | 51.1 (253) | 553 | 49.7 (275) |
| N+WSH | 1046 | 53.1 (555) | 365 | 51.8 (189) | 352 | 55.7 (196) | 329 | 51.7 (170) | 574 | 52.8 (303) | 472 | 53.4 (252) |
| Used antibiotics ≥2 times | | | | | | | | | | | | |
| C | 1143 | 13.3 (152) | 360 | 17.2 (62) | 394 | 11.7 (46) | 389 | 11.3 (44) | 571 | 12.8 (73) | 572 | 13.8 (79) |
| WSH | 1043 | 13.7 (143) | 366 | 14.2 (52) | 345 | 13.9 (48) | 332 | 13.0 (43) | 568 | 12.7 (72) | 475 | 15.0 (71) |
| N | 1048 | 11.6 (122) | 347 | 14.7 (51) | 358 | 12.0 (43) | 343 | 8.2 (28) | 495 | 12.7 (63) | 553 | 10.7 (59) |
| N+WSH | 1046 | 13.0 (136) | 365 | 14.8 (54) | 352 | 13.6 (48) | 329 | 10.3 (34) | 574 | 12.4 (71) | 472 | 13.8 (65) |
| Arm | N | Mean (SD) | N | Mean (SD) | N | Mean (SD) | N | Mean (SD) | N | Mean (SD) | N | Mean (SD) |
| Episodes of antibiotic use | | | | | | | | | | | | |
| C | 1143 | 0.68 (0.75) | 360 | 0.74 (0.85) | 394 | 0.65 (0.69) | 389 | 0.64 (0.69) | 571 | 0.67 (0.72) | 572 | 0.68 (0.77) |
| WSH | 1043 | 0.66 (0.77) | 366 | 0.68 (0.83) | 345 | 0.69 (0.74) | 332 | 0.61 (0.74) | 568 | 0.64 (0.76) | 475 | 0.69 (0.78) |
| N | 1048 | 0.65 (0.78) | 347 | 0.70 (0.93) | 358 | 0.68 (0.72) | 343 | 0.55 (0.65) | 495 | 0.66 (0.77) | 553 | 0.63 (0.79) |
| N+WSH | 1046 | 0.69 (0.78) | 365 | 0.71 (0.85) | 352 | 0.72 (0.78) | 329 | 0.63 (0.70) | 574 | 0.68 (0.77) | 472 | 0.71 (0.80) |
| Total days of antibiotic use | | | | | | | | | | | | |
| C | 1142 | 3.95 (12.1) | 359 | 5.61 (18.8) | 394 | 3.42 (9.11) | 389 | 2.95 (3.90) | 571 | 4.10 (12.7) | 571 | 3.80 (11.5) |
| WSH | 1043 | 4.14 (13.0) | 366 | 4.06 (10.9) | 345 | 5.37 (19.2) | 332 | 2.95 (4.12) | 568 | 4.53 (15.7) | 475 | 3.68 (8.72) |
| N | 1047 | 3.36 (8.45) | 347 | 2.90 (4.69) | 357 | 4.15 (12.0) | 343 | 2.99 (6.67) | 494 | 3.59 (9.60) | 553 | 3.15 (7.27) |
| N+WSH | 1046 | 3.71 (9.02) | 365 | 3.93 (9.96) | 352 | 3.56 (9.51) | 329 | 3.61 (7.22) | 574 | 3.60 (8.28) | 472 | 3.84 (9.86) |

C: Control; WSH: Water, sanitation, hygiene; N: Nutrition, N+WSH: Nutrition+ water, sanitation, hygiene; SD: Standard deviation.

**Table S8.** WSH, nutrition and N+WSH interventions vs. controls on antibiotic use within last 3 months among children in Bangladesh

| | | | Relative effects | | | | Absolute effects | | | |
|---|---|---|---|---|---|---|---|---|---|---|
| | | | Unadjusted | | Adjusted | | Unadjusted | | Adjusted | |
| Arm | n | % (n) | PR (95% CI) | p-value | PR (95% CI) | p-value | PD (95% CI) | p-value | PD (95% CI) | p-value |
| Used antibiotics ≥1 time | | | | | | | | | | |
| C | 1005 | 63.2 (635) | ref | -- | -- | -- | -- | -- | -- | -- |
| WSH | 1077 | 57.0 (614) | 0.90 (0.82, 0.99) | 0.03 | 0.90 (0.82, 0.99) | 0.03 | -0.06 (-0.12, -0.01) | 0.03 | -0.07 (-0.12, 0.03) | 0.03 |
| N | 1018 | 54.1 (551) | 0.86 (0.78, 0.94) | <0.001 | 0.84 (0.76, 0.92) | <0.001 | -0.09 (-0.14, -0.04) | <0.001 | -0.10 (-0.16, -0.05) | <0.001 |
| N+WSH | 1058 | 54.2 (573) | 0.86 (0.79, 0.93) | <0.001 | 0.86 (0.78, 0.93) | <0.001 | -0.09 (-0.14, -0.04) | <0.001 | -0.10 (-0.15, -0.04) | <0.001 |
| Used antibiotics ≥2 times | | | | | | | | | | |
| C | 1005 | 24.7 (248) | ref | -- | -- | -- | -- | -- | -- | -- |
| WSH | 1077 | 18.2 (196) | 0.74 (0.63, 0.87) | <0.001 | 0.73 (0.63, 0.85) | <0.001 | -0.06 (-0.10, -0.03) | <0.001 | -0.07 (-0.10, -0.04) | <0.001 |
| N | 1018 | 16.4 (167) | 0.66 (0.56, 0.79) | <0.001 | 0.64 (0.53, 0.78) | <0.001 | -0.08 (-0.12, -0.05) | <0.001 | -0.09 (-0.13, -0.05) | <0.001 |
| N+WSH | 1058 | 16.2 (171) | 0.65 (0.55, 0.78) | <0.001 | 0.66 (0.55, 0.79) | <0.001 | -0.09 (-0.12, -0.05) | <0.001 | -0.08 (-0.12, -0.05) | <0.001 |
| Arm | N | Mean (SD) | CR (95% CI) | p-value | CR (95% CI) | p-value | CD (95% CI) | p-value | CD (95% CI) | p-value |
| Episodes of antibiotic use | | | | | | | | | | |
| C | 1005 | 0.98 (0.98) | ref | -- | -- | -- | -- | -- | -- | -- |
| WSH | 1077 | 0.81 (0.88) | 0.83 (0.75, 0.92) | <0.001 | 0.82 (0.74. 0.90) | <0.001 | -0.17 (-0.26, -0.07) | <0.001 | -0.18 (-0.27, -0.09) | <0.001 |
| N | 1018 | 0.77 (0.90) | 0.79 (0.72, 0.88) | <0.001 | 0.77 (0.69, 0.86) | <0.001 | -0.20 (-0.29, -0.11) | <0.001 | -0.22 (-0.32, -0.13) | <0.001 |
| N+WSH | 1058 | 0.77 (0.89) | 0.79 (0.71, 0.87) | <0.001 | 0.78 (0.71, 0.87) | <0.001 | -0.21 (-0.29, -0.12) | <0.001 | -0.21 (-0.30, -0.12) | <0.001 |
| Total days of antibiotic use | | | | | | | | | | |
| C | 995 | 5.06 (5.81) | ref | -- | -- | -- | -- | -- | -- | -- |
| WSH | 1075 | 4.11 (5.05) | 0.81 (0.74, 0.90) | <0.001 | 0.81 (0.73, 0.89) | <0.001 | -0.94 (-1.41, -0.48) | <0.001 | -1.02 (-1.49, -0.54) | <0.001 |
| N | 1016 | 4.02 (5.30) | 0.79 (0.71, 0.89) | <0.001 | 0.75 (0.67, 0.85) | <0.001 | -1.05 (-1.54, -0.55) | <0.001 | -1.19 (-1.74, -0.65) | <0.001 |
| N+WSH | 1055 | 4.05 (5.39) | 0.80 (0.72, 0.89) | <0.001 | 0.80 (0.71, 0.89) | <0.001 | -1.01 (-1.51, -0.51) | <0.001 | -1.06 (-1.60, -0.53) | <0.001 |

PR: Prevalence ratio (for binary outcomes), CR: Count ratio (for count outcomes); PD: Prevalence difference (for binary outcomes); CD: Count difference (for count outcomes); CI: Confidence interval; C: Control; WSH: Water, sanitation, hygiene; N: Nutrition, N+WSH: Nutrition+ water, sanitation, hygiene.

**Table S9.** Combined N+WSH intervention vs. WSH and nutrition interventions on antibiotic use within last 3 months among children in Bangladesh

| | Relative effects | | | | Absolute effects | | | |
|---|---|---|---|---|---|---|---|---|
| | Unadjusted | | Adjusted | | Unadjusted | | Adjusted | |
| Arm | PR (95% CI) | p-value | PR (95% CI) | p-value | PD (95% CI) | p-value | PD (95% CI) | p-value |
| Used antibiotics ≥1 time | | | | | | | | |
| N+WSH vs. WSH | 0.95 (0.87, 1.03) | 0.22 | 0.95 (0.88, 1.04) | 0.26 | -0.03 (-0.07, 0.02) | 0.22 | -0.02 (-0.07, 0.02) | 0.35 |
| N+WSH vs. N | 1.00 (0.91, 1.10) | 0.99 | 1.01 (0.92, 1.12) | 0.77 | 0.00 (-0.05, 0.05) | 0.99 | 0.01 (-0.05, 0.06) | 0.77 |
| Used antibiotics ≥2 times | | | | | | | | |
| N+WSH vs. WSH | 0.89 (0.74, 1.06) | 0.19 | 0.91 (0.76, 1.09) | 0.32 | -0.02 (-0.05, 0.01) | 0.19 | -0.01 (-0.04, 0.01) | 0.40 |
| N+WSH vs. N | 0.99 (0.83, 1.17) | 0.87 | 1.05 (0.88, 1.25) | 0.62 | -0.00 (-0.03, 0.03) | 0.87 | 0.01 (-0.02, 0.04) | 0.59 |
| Arm | CR (95% CI) | p-value | CR (95% CI) | p-value | CD (95% CI) | p-value | CD (95% CI) | p-value |
| Episodes of antibiotic use | | | | | | | | |
| N+WSH vs. WSH | 0.95 (0.85, 1.06) | 0.35 | 0.98 (0.88, 1.09) | 0.68 | -0.04 (-0.13, 0.04) | 0.35 | -0.02 (-0.10, 0.07) | 0.67 |
| N+WSH vs. N | 0.99 (0.89, 1.10) | 0.87 | 1.02 (0.91, 1.14) | 0.76 | -0.01 (-0.09, 0.08) | 0.87 | 0.01 (-0.07, 0.10) | 0.75 |
| Total days of antibiotic use | | | | | | | | |
| N+WSH vs. WSH | 0.98 (0.87, 1.11) | 0.79 | 0.99 (0.87, 1.11) | 0.83 | -0.07 (-0.55, 0.41) | 0.79 | 0.04 (-0.45, 0.53) | 0.87 |
| N+WSH vs. N | 1.01 (0.90, 1.13) | 0.88 | 1.05 (0.93, 1.18) | 0.41 | 0.03 (-0.42, 0.49) | 0.88 | 0.17 (-0.30, 0.63) | 0.48 |

PR: Prevalence ratio (for binary outcomes), CR: Count ratio (for count outcomes); PD: Prevalence difference (for binary outcomes); CD: Count difference (for count outcomes); CI: Confidence interval; WSH: Water, sanitation, hygiene; N: Nutrition, N+WSH: Nutrition+ water, sanitation, hygiene.

**Table S10.** WSH, nutrition and N+WSH interventions vs. controls on antibiotic use within last 3 months among children in Kenya

| | | | Relative effects | | | | Absolute effects | | | |
|---|---|---|---|---|---|---|---|---|---|---|
| | | | Unadjusted | | Adjusted | | Unadjusted | | Adjusted | |
| Arm | N | % (n) | PR (95% CI) | p-value | PR (95% CI) | p-value | PD (95% CI) | p-value | PD (95% CI) | p-value |
| Used antibiotics ≥1 time | | | | | | | | | | |
| C | 1143 | 52.6 (601) | ref | -- | -- | -- | -- | -- | -- | -- |
| WSH | 1043 | 50.3 (525) | 0.96 (0.86, 1.06) | 0.41 | 0.97 (0.88, 1.07) | 0.55 | -0.02 (-0.08, 0.03) | 0.41 | -0.02 (-0.07, 0.04) | 0.53 |
| N | 1048 | 50.4 (528) | 0.96 (0.88, 1.04) | 0.33 | 0.95 (0.87, 1.03) | 0.19 | -0.02 (-0.07, 0.02) | 0.33 | -0.03 (-0.07, 0.02) | 0.22 |
| N+WSH | 1046 | 53.1 (555) | 1.01 (0.92, 1.10) | 0.84 | 1.00 (0.91, 1.09) | 0.99 | 0.00 (-0.04, 0.05) | 0.84 | 0.00 (-0.05, 0.05) | 0.97 |
| Used antibiotics ≥2 times | | | | | | | | | | |
| C | 1143 | 13.3 (152) | ref | -- | -- | -- | -- | -- | -- | -- |
| WSH | 1043 | 13.7 (143) | 1.03 (0.80. 1.32) | 0.81 | 1.03 (0.79, 1.35) | 0.81 | 0.00 (-0.03, 0.04) | 0.81 | 0.00 (-0.03, 0.04) | 0.80 |
| N | 1048 | 11.6 (122) | 0.88 (0.68, 1.12) | 0.30 | 0.88 (0.68, 1.14) | 0.33 | -0.02 (-0.05, 0.01) | 0.30 | -0.02 (-0.05, 0.02) | 0.33 |
| N+WSH | 1046 | 13.0 (136) | 0.98 (0.78, 1.22) | 0.84 | 0.94 (0.74, 1.19) | 0.61 | -0.00 (-0.03, 0.03) | 0.84 | -0.01 (-0.04, 0.02) | 0.61 |
| Arm | N | Mean (SD) | CR (95% CI) | p-value | CR (95% CI) | p-value | CD (95% CI) | p-value | CD (95% CI) | p-value |
| Episodes of antibiotic use | | | | | | | | | | |
| C | 1143 | 0.68 (0.75) | ref | -- | -- | -- | -- | -- | -- | -- |
| WSH | 1043 | 0.66 (0.77) | 0.98 (0.87, 1.10) | 0.77 | 0.99 (0.89, 1.12) | 0.92 | -0.01 (-0.09, 0.07) | 0.77 | -0.00 (-0.08, 0.07) | 0.93 |
| N | 1048 | 0.65 (0.78) | 0.96 (0.85, 1.07) | 0.45 | 0.96 (0.86, 1.07) | 0.41 | -0.03 (-0.10, 0.05) | 0.44 | -0.03 (-0.10, 0.04) | 0.40 |
| N+WSH | 1046 | 0.69 (0.78) | 1.02 (0.92, 1.14) | 0.66 | 1.00 (0.90, 1.12) | 0.90 | 0.02 (-0.06, 0.09) | 0.67 | 0.00 (-0.07, 0.08) | 0.90 |
| Total days of antibiotic use | | | | | | | | | | |
| C | 1142 | 3.95 (12.1) | ref | -- | -- | -- | -- | -- | -- | -- |
| WSH | 1043 | 4.14 (13.0) | 1.05 (0.71, 1.54) | 0.81 | 1.00 (0.73, 1.37) | 0.99 | 0.19 (-1.37, 1.76) | 0.81 | 0.25 (-1.37, 1.88) | 0.76 |
| N | 1047 | 3.36 (8.45) | 0.85 (0.65, 1.12) | 0.24 | 0.83 (0.63, 1.09) | 0.17 | -0.59 (-1.61, 0.43) | 0.25 | -0.68 (-1.68, 0.32) | 0.18 |
| N+WSH | 1046 | 3.71 (9.02) | 0.94 (0.76, 1.17) | 0.57 | 0.95 (0.77, 1.16) | 0.60 | -0.24 (-1.08, 0.60) | 0.58 | -0.29 (-1.16, 0.58) | 0.51 |

PR: Prevalence ratio (for binary outcomes), CR: Count ratio (for count outcomes); PD: Prevalence difference (for binary outcomes); CD: Count difference (for count outcomes); CI: Confidence interval; C: Control; WSH: Water, sanitation, hygiene; N: Nutrition, N+WSH: Nutrition+ water, sanitation, hygiene.

**Table S11.** N+WSH intervention vs. WSH and nutrition interventions on antibiotic use within last 3 months among children in Kenya

| | Relative effects | | | | Absolute effects | | | |
|---|---|---|---|---|---|---|---|---|
| | Unadjusted | | Adjusted | | Unadjusted | | Adjusted | |
| Arm | PR (95% CI) | p-value | PR (95% CI) | p-value | PD (95% CI) | p-value | PD (95% CI) | p-value |
| Used antibiotics ≥1 time | | | | | | | | |
| N+WSH vs. WSH | 1.05 (0.95, 1.17) | 0.33 | 1.04 (0.93, 1.15) | 0.50 | 0.03 (-0.03, 0.08) | 0.33 | 0.02 (-0.04, 0.07) | 0.54 |
| N+WSH vs. N | 1.05 (0.95, 1.17) | 0.32 | 1.05 (0.95, 1.16) | 0.36 | 0.03 (-0.03, 0.08) | 0.32 | 0.03 (-0.03, 0.08) | 0.35 |
| Used antibiotics ≥2 times | | | | | | | | |
| N+WSH vs. WSH | 0.95 (0.71, 1.27) | 0.72 | 0.95 (0.70, 1.28) | 0.72 | -0.01 (-0.05, 0.03) | 0.73 | -0.01 (-0.05, 0.03) | 0.73 |
| N+WSH vs. N | 1.12 (0.82, 1.52) | 0.48 | 1.12 (0.81, 1.54) | 0.49 | 0.01 (-0.02, 0.05) | 0.49 | 0.01 (-0.03, 0.05) | 0.51 |
| Arm | CR (95% CI) | p-value | CR (95% CI) | p-value | CD (95% CI) | p-value | CD (95% CI) | p-value |
| Episodes of antibiotic use | | | | | | | | |
| N+WSH vs. WSH | 1.04 (0.91, 1.19) | 0.56 | 1.02 (0.89, 1.17) | 0.72 | 0.03 (-0.06, 0.12) | 0.56 | 0.02 (-0.08, 0.11) | 0.73 |
| N+WSH vs. N | 1.07 (0.93, 1.23) | 0.34 | 1.08 (0.93, 1.24) | 0.31 | 0.05 (-0.05, 0.14) | 0.34 | 0.05 (-0.05, 0.14) | 0.33 |
| Total days of antibiotic use | | | | | | | | |
| N+WSH vs. WSH | 0.90 (0.64, 1.26) | 0.52 | 0.95 (0.72, 1.25) | 0.70 | -0.43 (-1.81, 0.94) | 0.53 | -0.48 (-1.86, 0.89) | 0.49 |
| N+WSH vs. N | 1.10 (0.87, 1.40) | 0.41 | 1.19 (0.95, 1.49) | 0.13 | 0.35 (-0.48, 1.18) | 0.41 | 0.50 (-0.33, 1.33) | 0.24 |

PR: Prevalence ratio (for binary outcomes), CR: Count ratio (for count outcomes); PD: Prevalence difference (for binary outcomes); CD: Count difference (for count outcomes); CI: Confidence interval; WSH: Water, sanitation, hygiene; N: Nutrition, N+WSH: Nutrition+ water, sanitation, hygiene.

**Table S12.** Multiplicative interaction by age and sex for unadjusted intervention effects on antibiotic use within last 3 months, Bangladesh

| | WSH vs. C | | N vs. C | | N+WSH vs. C | | N+WSH vs. WSH | | N+WSH vs. N | |
|---|---|---|---|---|---|---|---|---|---|---|
| | PR (95% CI) | p-value[a] | PR (95% CI) | p-value[a] | PR (95% CI) | p-value[a] | PR (95% CI) | p-value[a] | PR (95% CI) | p-value[a] |
| Used antibiotics ≥1 time | | | | | | | | | | |
| All obs | **0.90 (0.82, 0.99)** | | **0.86 (0.78, 0.94)** | | **0.86 (0.79, 0.93)** | | 0.95 (0.87, 1.03) | | 1.00 (0.91, 1.10) | |
| Age 3 mo | 0.84 (0.70, 1.02) | -- | **0.76 (0.63, 0.93)** | -- | **0.67 (0.54, 0.82)** | -- | **0.79 (0.62, 1.00)** | -- | 0.87 (0.67, 1.13) | -- |
| Age 14 mo | 0.94 (0.85, 1.04) | 0.26 | 0.91 (0.82, 1.00) | 0.10 * | 0.95 (0.86, 1.05) | <0.001 * | 1.01 (0.92, 1.11) | 0.06 * | 1.05 (0.94, 1.17) | 0.15 * |
| Age 28 mo | 0.90 (0.78, 1.03) | 0.57 | **0.85 (0.73, 1.00)** | 0.39 | 0.88 (0.76, 1.00) | 0.04 * | 0.98 (0.87, 1.10) | 0.09 * | 1.02 (0.89, 1.18) | 0.30 |
| Female | **0.88 (0.77, 1.00)** | -- | **0.80 (0.70, 0.90)** | -- | **0.80 (0.71, 0.90)** | -- | 0.91 (0.80, 1.03) | -- | 1.00 (0.87, 1.15) | -- |
| Male | 0.92 (0.83, 1.03) | 0.49 | 0.91 (0.81, 1.02) | 0.09 * | 0.92 (0.83, 1.03) | 0.05 * | 1.00 (0.90, 1.11) | 0.24 | 1.01 (0.91, 1.13) | 0.88 |
| Used antibiotics ≥2 times | | | | | | | | | | |
| All obs | **0.74 (0.63, 0.87)** | | **0.66 (0.56, 0.79)** | | **0.65 (0.55, 0.78)** | | 0.89 (0.74, 1.06) | | 0.99 (0.83, 1.17) | |
| Age 3 mo | **0.57 (0.37, 0.86)** | -- | **0.51 (0.33, 0.78)** | -- | **0.48 (0.33, 0.69)** | -- | 0.85 (0.54, 1.32) | -- | 0.94 (0.61, 1.44) | -- |
| Age 14 mo | **0.74 (0.60, 0.90)** | 0.26 | **0.70 (0.57, 0.86)** | 0.18 * | **0.77 (0.64, 0.94)** | 0.02 * | 1.05 (0.84, 1.31) | 0.40 | 1.10 (0.90. 1.36) | 0.52 |
| Age 28 mo | 0.99 (0.75, 1.32) | 0.04 * | 0.79 (0.55, 1.13) | 0.12 * | **0.64 (0.42, 0.98)** | 0.29 | **0.65 (0.45, 0.93)** | 0.38 | 0.81 (0.53, 1.26) | 0.64 |
| Female | **0.72 (0.55, 0.93)** | -- | **0.71 (0.56, 0.90)** | -- | **0.66 (0.52, 0.83)** | -- | 0.91 (0.69, 1.20) | -- | 0.93 (0.70, 1.22) | -- |
| Male | **0.74 (0.60, 0.91)** | 0.85 | **0.63 (0.50, 0.79)** | 0.47 | **0.66 (0.51, 0.85)** | 0.97 | 0.89 (0.70, 1.13) | 0.89 | 1.06 (0.81, 1.37) | 0.54 |
| | CR (95% CI) | p-value[a] | CR (95% CI) | p-value[a] | CR (95% CI) | p-value[a] | CR (95% CI) | p-value[a] | CR (95% CI) | p-value[a] |
| Episodes of antibiotic use | | | | | | | | | | |
| All obs | **0.83 (0.75, 0.92)** | | **0.79 (0.72, 0.88)** | | **0.79 (0.71, 0.87)** | | 0.95 (0.85, 1.06) | | 0.99 (0.89, 1.10) | |
| Age 3 mo | **0.72 (0.57, 0.92)** | -- | **0.65 (0.50, 0.84)** | -- | **0.62 (0.49, 0.80)** | -- | 0.86 (0.67, 1.11) | -- | 0.96 (0.74, 1.25) | -- |
| Age 14 mo | **0.85 (0.75, 0.96)** | 0.19 * | **0.85 (0.75, 0.96)** | 0.05 * | **0.88 (0.79, 0.99)** | 0.01 * | 1.03 (0.90, 1.18) | 0.18 * | 1.04 (0.91, 1.19) | 0.59 |
| Age 28 mo | 0.89 (0.77, 1.03) | 0.14 * | **0.84 (0.71,1.00)** | 0.11 * | **0.80 (0.68, 0.94)** | 0.10 * | 0.90 (0.78, 1.04) | 0.75 | 0.95 (0.81, 1.13) | 0.97 |
| Female | **0.80 (0.69, 0.93)** | -- | **0.76 (0.66, 0.88)** | -- | **0.76 (0.67, 0.86)** | -- | 0.94 (0.81, 1.09) | -- | 1.00 (0.86, 1.16) | -- |
| Male | **0.84 (0.73, 0.97)** | 0.65 | **0.82 (0.71, 0.94)** | 0.48 | **0.82 (0.71, 0.95)** | 0.43 | 0.97 (0.84, 1.13) | 0.75 | 1.00 (0.87, 1.16) | 0.94 |
| Total days of antibiotic use | | | | | | | | | | |
| All obs | **0.81 (0.74, 0.90)** | | **0.79 (0.71, 0.89)** | | **0.80 (0.72, 0.89)** | | 0.98 (0.87, 1.11) | | 1.01 (0.90, 1.13) | |
| Age 3 mo | **0.74 (0.58, 0.94)** | -- | **0.63 (0.48, 0.83)** | -- | **0.68 (0.52, 0.89)** | -- | 0.92 (0.69, 1.22) | -- | 1.08 (0.81, 1.46) | -- |
| Age 14 mo | **0.82 (0.72, 0.93)** | 0.44 | **0.85 (0.75, 0.97)** | 0.05 * | 0.91 (0.79, 1.05) | 0.04 * | 1.11 (0.96, 1.29) | 0.19 * | 1.07 (0.93, 1.23) | 0.93 |
| Age 28 mo | 0.88 (0.74, 1.05) | 0.30 | 0.85 (0.70, 1.03) | 0.09 * | **0.76 (0.64, 0.90)** | 0.54 * | 0.86 (0.73, 1.01) | 0.70 | 0.89 (0.74, 1.07) | 0.29 |
| Female | **0.80 (0.67, 0.95)** | -- | **0.77 (0.64, 0.92)** | -- | **0.76 (0.66, 0.89)** | -- | 0.95 (0.82, 1.12) | -- | 0.99 (0.84, 1.18) | -- |
| Male | **0.82 (0.72, 0.93)** | 0.87 | **0.81 (0.70, 0.94)** | 0.68 | **0.84 (0.72, 0.98)** | 0.36 | 1.03 (0.87, 1.22) | 0.52 | 1.04 (0.88, 1.23) | 0.72 |

PR: Prevalence ratio (for binary outcomes), CR: Count ratio (for count outcomes); CI: Confidence interval; C: Control; WSH: Water, sanitation, hygiene; N: Nutrition, N+WSH: Nutrition+ water, sanitation, hygiene.

Bolded values refer to estimates with a p-value ≤0.05.

[a] p-value for interaction term between study arm and subgroup variable.

* Interaction p-values <0.20 indicate significant effect modification.

**Table S13.** Additive interaction by age and sex for unadjusted intervention effects on antibiotic use within last 3 months, Bangladesh

| | WSH vs. C | | N vs. C | | N+WSH vs. C | | N+WSH vs. WSH | | N+WSH vs. N | |
|---|---|---|---|---|---|---|---|---|---|---|
| | PD (95% CI) | p-value[a] | PD (95% CI) | p-value[a] | PD (95% CI) | p-value[a] | PD (95% CI) | p-value[a] | PD (95% CI) | p-value[a] |
| Used antibiotics ≥1 time | | | | | | | | | | |
| All obs | **-0.06 (-0.12, -0.01)** | | **-0.09 (-0.14, -0.04)** | | **-0.09 (-0.14, -0.04)** | | -0.03 (-0.07, 0.02) | | 0.00 (-0.05, 0.05) | |
| Age 3 mo | -0.08 (-0.17, 0.01) | -- | **-0.13 (-0.22, -0.04)** | -- | **-0.18 (-0.27, -0.09)** | -- | **-0.10 (-0.19, 0.00)** | -- | -0.05 (-0.15, 0.05) | -- |
| Age 14 mo | -0.04 (-0.12, 0.03) | 0.43 | -0.07 (-0.14, 0.00) | 0.28 | -0.04 (-0.11, 0.04) | 0.01 * | 0.01 (-0.06, 0.08) | 0.09 * | 0.03 (-0.04, 0.11) | 0.12 * |
| Age 28 mo | -0.06 (-0.14, 0.02) | 0.64 | **-0.08 (-0.16, -0.00)** | 0.49 | -0.07 (-0.15, 0.00) | 0.08 * | -0.01 (-0.07, 0.04) | 0.12 * | 0.01 (-0.06, 0.08) | 0.32 |
| Female | **-0.08 (-0.15, -0.00)** | -- | **-0.13 (-0.19, -0.07)** | -- | **-0.13 (-0.19, -0.06)** | -- | -0.05 (-0.12, 0.02) | -- | 0.00 (-0.07, 0.07) | -- |
| Male | -0.05 (-0.12, 0.02) | 0.54 | -0.06 (-0.13, 0.01) | 0.12 * | -0.05 (-0.12, 0.02) | 0.06 * | 0.00 (-0.06, 0.06) | 0.27 | 0.01 (-0.06, 0.07) | 0.87 |
| Used antibiotics ≥2 times | | | | | | | | | | |
| All obs | **-0.06 (-0.10, -0.03)** | | **-0.08 (-0.12, -0.05)** | | **-0.09 (-0.12, -0.05)** | | -0.02 (-0.05, 0.01) | | 0.00 (-0.03, 0.03) | |
| Age 3 mo | **-0.11 (-0.18, -0.03)** | -- | **-0.12 (-0.19, -0.05)** | -- | **-0.13 (-0.19, -0.06)** | -- | -0.02 (-0.08, 0.04) | -- | -0.01 (-0.06, 0.05) | -- |
| Age 14 mo | **-0.09 (-0.15, -0.03)** | 0.73 | **-0.10 (-0.16, -0.05)** | 0.68 | **-0.08 (-0.13, -0.02)** | 0.22 | 0.01 (-0.04, 0.07) | 0.42 | 0.03 (-0.03, 0.08) | 0.41 |
| Age 28 mo | -0.00 (-0.04, 0.04) | 0.02 * | -0.03 (-0.07, 0.01) | 0.03 * | **-0.05 (-0.10, -0.00)** | 0.04 * | **-0.05 (-0.09, -0.01)** | 0.47 | -0.02 (-0.06, 0.02) | 0.71 |
| Female | **-0.06 (-0.11, -0.01)** | -- | **-0.06 (-0.11, -0.02)** | -- | **-0.08 (-0.12, -0.03)** | -- | -0.01 (-0.06, 0.03) | -- | -0.01 (-0.05, 0.03) | -- |
| Male | **-0.07 (-0.12, -0.02)** | 0.82 | **-0.10 (-0.15, -0.05)** | 0.26 | **-0.09 (-0.15, -0.04)** | 0.64 | -0.02 (-0.07, 0.02) | 0.78 | 0.01 (-0.04, 0.06) | 0.55 |
| | CD (95% CI) | p-value[a] | CD (95% CI) | p-value[a] | CD (95% CI) | p-value[a] | CD (95% CI) | p-value[a] | CD (95% CI) | p-value[a] |
| Episodes of antibiotic use | | | | | | | | | | |
| All obs | -0.17 (-0.26, -0.07) | <0.001 | -0.20 (-0.29, -0.11) | <0.001 | -0.21 (-0.29, -0.12) | <0.001 | -0.04 (-0.13, 0.04) | 0.35 | -0.01 (-0.09, 0.08) | 0.87 |
| Age 3 mo | **-0.25 (-0.43, -0.07)** | -- | **-0.32 (-0.50, -0.14)** | -- | **-0.34 (-0.51, -0.17)** | -- | -0.06 (-0.13, 0.02) | -- | -0.02 (-0.10, 0.05) | -- |
| Age 14 mo | **-0.18 (-0.32, -0.04)** | 0.52 | **-0.19 (-0.32, -0.05)** | 0.23 | **-0.15 (-0.28, -0.02)** | 0.05 * | -0.03 (-0.10, 0.04) | 0.64 | 0.02 (-0.05, 0.09) | 0.37 |
| Age 28 mo | -0.08 (-0.19, 0.03) | 0.11 * | **-0.12 (-0.24, -0.00)** | 0.09 * | **-0.15 (-0.26, -0.04)** | 0.08 * | -0.00 (-0.05, 0.05) | 0.23 | -0.02 (-0.07, 0.03) | 0.95 |
| Female | **-0.18 (-0.30, -0.06)** | -- | **-0.22 (-0.33, -0.11)** | -- | **-0.22 (-0.32, -0.12)** | -- | -0.04 (-0.15, 0.06) | -- | -0.00 (-0.11, 0.10) | -- |
| Male | **-0.16 (-0.29, -0.03)** | 0.84 | **-0.19 (-0.32, -0.06)** | 0.70 | **-0.18 (-0.32, -0.05)** | 0.66 | -0.02 (-0.15, 0.10) | 0.81 | 0.00 (-0.12, 0.13) | 0.94 |
| Total days of antibiotic use | | | | | | | | | | |
| All obs | -0.94 (-1.41, -0.48) | <0.001 | -1.05 (-1.54, -0.55) | <0.001 | -1.01 (-1.51, -0.51) | <0.001 | -0.07 (-0.55, 0.41) | 0.79 | 0.03 (-0.42, 0.49) | 0.88 |
| Age 3 mo | **-1.23 (-2.20, -0.26)** | -- | **-1.77 (-2.75, -0.79)** | -- | **-1.52 (-2.55, -0.49)** | -- | -0.09 (-0.25, 0.06) | -- | -0.02 (-0.17, 0.13) | |
| Age 14 mo | **-1.14 (-1.86, -0.43)** | 0.87 | **-0.93 (-1.69, -0.18)** | 0.17 * | -0.57 (-1.41, 0.27) | 0.12 * | 0.04 (-0.11, 0.18) | 0.20 | 0.04 (-0.11, 0.19) | 0.52 |
| Age 28 mo | -0.49 (-1.16, 0.18) | 0.25 | -0.60 (-1.32, 0.12) | 0.07 * | **-0.98 (-1.60, -0.36)** | 0.41 | -0.07 (-0.16, 0.02) | 0.80 | -0.03 (-0.13, 0.08) | 0.95 |
| Female | **-0.95 (-1.72, -0.18)** | -- | **-1.10 (-1.85, -0.35)** | -- | **-1.12 (-1.75, -0.49)** | -- | -0.17 (-0.76, 0.42) | -- | -0.02 (-0.65, 0.60) | -- |
| Male | **-0.98 (-1.63, -0.33)** | 0.96 | **-1.03 (-1.74, -0.32)** | 0.90 | **-0.85 (-1.60, -0.11)** | 0.58 | 0.13 (-0.62, 0.88) | 0.54 | 0.17 (-0.55, 0.90) | 0.70 |

PD: Prevalence difference (for binary outcomes); CD: Count difference (for count outcomes); CI: Confidence interval; C: Control; WSH: Water, sanitation, hygiene; N: Nutrition, N+WSH: Nutrition+ water, sanitation, hygiene.

Bolded values refer to estimates with a p-value ≤0.05.

[a] p-value for interaction term between study arm and subgroup variable.

* Interaction p-values <0.20 indicate significant effect modification.

**Table S14.** Multiplicative interaction by age and sex for unadjusted intervention effects on antibiotic use within last 3 months, Kenya

| | WSH vs. C | | N vs. C | | N+WSH vs. C | | N+WSH vs. WSH | | N+WSH vs. N | |
|---|---|---|---|---|---|---|---|---|---|---|
| | PR (95% CI) | p-value [a] | PR (95% CI) | p-value [a] | PR (95% CI) | p-value [a] | PR (95% CI) | p-value [a] | PR (95% CI) | p-value [a] |
| Used antibiotics ≥1 time | | | | | | | | | | |
| All obs | 0.96 (0.86, 1.06) | 0.41 | 0.96 (0.88, 1.04) | 0.33 | 1.01 (0.92, 1.10) | 0.84 | 1.05 (0.95, 1.17) | 0.33 | 1.05 (0.95, 1.17) | 0.32 |
| Age 6 mo | 0.95 (0.83, 1.08) | -- | 0.94 (0.82, 1.08) | -- | 0.99 (0.84, 1.16) | -- | 1.04 (0.87, 1.25) | -- | 1.06 (0.89, 1.24) | -- |
| Age 17 mo | 1.01 (0.85, 1.20) | 0.57 | 1.03 (0.90, 1.17) | 0.34 | 1.04 (0.91, 1.20) | 0.63 | 1.03 (0.91, 1.18) | 0.93 | 1.02 (0.88, 1.17) | 0.75 |
| Age 22 mo | 0.91 (0.76, 1.09) | 0.74 | 0.90 (0.78, 1.04) | 0.70 | 1.00 (0.85, 1.16) | 0.94 | 1.09 (0.92, 1.30) | 0.70 | 1.10 (0.94, 1.29) | 0.67 |
| Female | 0.93 (0.81, 1.06) | -- | 0.96 (0.86, 1.07) | -- | 0.99 (0.85, 1.15) | -- | 1.07 (0.93, 1.22) | -- | 1.03 (0.89, 1.20) | -- |
| Male | 0.99 (0.86, 1.14) | 0.50 | 0.96 (0.85, 1.08) | 0.98 | 1.03 (0.92, 1.15) | 0.72 | 1.04 (0.89, 1.21) | 0.79 | 1.07 (0.96, 1.21) | 0.67 |
| Used antibiotics ≥2 times | | | | | | | | | | |
| All obs | 1.03 (0.80. 1.32) | 0.81 | 0.88 (0.68, 1.12) | 0.30 | 0.98 (0.78, 1.22) | 0.84 | 0.95 (0.71, 1.27) | 0.72 | 1.12 (0.82, 1.52) | 0.48 |
| Age 6 mo | 0.83 (0.60, 1.14) | -- | 0.85 (0.59, 1.23) | -- | 0.86 (0.59, 1.26) | -- | 1.04 (0.67, 1.62) | -- | 1.01 (0.65, 1.56) | -- |
| Age 17 mo | 1.19 (0.77, 1.84) | 0.20 | 1.03 (0.66, 1.60) | 0.52 | 1.17 (0.79, 1.72) | 0.31 | 0.98 (0.62, 1.55) | 0.85 | 1.14 (0.76, 1.71) | 0.65 |
| Age 22 mo | 1.15 (0.75, 1.75) | 0.18 * | 0.72 (0.48, 1.08) | 0.54 | 0.91 (0.59, 1.41) | 0.84 | 0.80 (0.48, 1.32) | 0.39 | 1.27 (0.80, 1.99) | 0.41 |
| Female | 0.99 (0.74, 1.33) | -- | 1.00 (0.72, 1.37) | -- | 0.97 (0.70, 1.34) | -- | 0.98 (0.71, 1.35) | -- | 0.97 (0.66, 1.43) | -- |
| Male | 1.08 (0.75, 1.56) | 0.70 | 0.77 (0.53, 1.13) | 0.31 | 1.00 (0.73, 1.37) | 0.90 | 0.92 (0.59, 1.44) | 0.83 | 1.29 (0.85, 1.96) | 0.27 |
| | CR (95% CI) | p-value [a] | CR (95% CI) | p-value [a] | CR (95% CI) | p-value [a] | CR (95% CI) | p-value [a] | CR (95% CI) | p-value [a] |
| Episodes of antibiotic use | | | | | | | | | | |
| All obs | 0.98 (0.87, 1.10) | 0.77 | 0.95 (0.85, 1.07) | 0.45 | 1.02 (0.92, 1.14) | 0.66 | 1.04 (0.91, 1.19) | 0.56 | 1.07 (0.93, 1.23) | 0.34 |
| Age 6 mo | 0.92 (0.79, 1.08) | -- | 0.94 (0.78, 1.14) | -- | 0.96 (0.79, 1.17) | -- | 1.05 (0.83, 1.32) | -- | 1.02 (0.82, 1.28) | -- |
| Age 17 mo | 1.06 (0.87, 1.29) | 0.31 | 1.05 (0.89, 1.22) | 0.42 | 1.11 (0.95, 1.29) | 0.31 | 1.05 (0.87, 1.26) | 0.99 | 1.06 (0.89, 1.26) | 0.77 |
| Age 22 mo | 0.96 (0.79, 1.18) | 0.72 | 0.87 (0.75, 1.01) | 0.45 | 1.00 (0.84, 1.18) | 0.83 | 1.03 (0.83, 1.27) | 0.91 | 1.14 (0.96, 1.36) | 0.37 |
| Female | 0.96 (0.83, 1.11) | -- | 0.99 (0.86, 1.13) | -- | 1.01 (0.86, 1.19) | -- | 1.05 (0.90, 1.24) | -- | 1.03 (0.86, 1.23) | -- |
| Male | 1.01 (0.85, 1.19) | 0.63 | 0.93 (0.79, 1.09) | 0.53 | 1.04 (0.91, 1.19) | 0.83 | 1.03 (0.84, 1.26) | 0.84 | 1.12 (0.95, 1.32) | 0.42 |
| Total days of antibiotic use | | | | | | | | | | |
| All obs | 1.05 (0.71, 1.54) | 0.81 | 0.85 (0.65, 1.12) | 0.24 | 0.94 (0.76, 1.17) | 0.57 | 0.90 (0.64, 1.27) | 0.52 | 1.10 (0.87, 1.40) | 0.41 |
| Age 6 mo | 0.72 (0.42, 1.25) | -- | **0.52 (0.35, 0.77)** | -- | 0.70 (0.43, 1.14) | -- | 0.97 (0.62, 1.51) | -- | 1.36 (0.97, 1.90) | -- |
| Age 17 mo | 1.57 (0.79, 3.13) | 0.08 * | 1.21 (0.75, 1.95) | 0.01 * | 1.04 (0.74, 1.46) | 0.26 | 0.66 (0.34, 1.30) | 0.33 | 0.86 (0.56, 1.33) | 0.12 * |
| Age 22 mo | 1.00 (0.78, 1.28) | 0.27 | 1.01 (0.75, 1.37) | 0.01 * | 1.22 (0.91, 1.65) | 0.06 * | 1.22 (0.92, 1.63) | 0.42 | 1.21 (0.87, 1.67) | 0.59 |
| Female | 1.10 (0.67, 1.83) | -- | 0.88 (0.62, 1.24) | -- | 0.88 (0.64, 1.20) | -- | 0.80 (0.50, 1.26) | -- | 1.00 (0.72, 1.39) | -- |
| Male | 0.97 (0.67, 1.40) | 0.60 | 0.83 (0.57, 1.21) | 0.83 | 1.01 (0.73, 1.39) | 0.55 | 1.04 (0.74, 1.46) | 0.27 | 1.22 (0.88, 1.70) | 0.40 |

PR: Prevalence ratio (for binary outcomes), CR: Count ratio (for count outcomes); CI: Confidence interval; C: Control; WSH: Water, sanitation, hygiene; N: Nutrition, N+WSH: Nutrition+ water, sanitation, hygiene.

Bolded values refer to estimates with a p-value ≤0.05.

[a] p-value for interaction term between study arm and subgroup variable.

* Interaction p-values <0.20 indicate significant effect modification.

**Table S15.** Additive interaction by age and sex for unadjusted intervention effects on antibiotic use within last 3 months, Kenya

| | WSH vs. C | | N vs. C | | N+WSH vs. C | | N+WSH vs. WSH | | N+WSH vs. N | |
|---|---|---|---|---|---|---|---|---|---|---|
| | PD (95% CI) | p-value[a] | PD (95% CI) | p-value[a] | PD (95% CI) | p-value[a] | PD (95% CI) | p-value[a] | PD (95% CI) | p-value[a] |
| Used antibiotics ≥1 time | | | | | | | | | | |
| All obs | -0.02 (-0.08, 0.03) | 0.41 | -0.02 (-0.07, 0.02) | 0.33 | 0.00 (-0.05, 0.05) | 0.84 | 0.03 (-0.03, 0.08) | 0.33 | 0.03 (-0.03, 0.08) | 0.32 |
| Age 6 mo | -0.03 (-0.10, 0.04) | -- | -0.03 (-0.10, 0.04) | -- | -0.01 (-0.09, 0.08) | -- | 0.02 (-0.07, 0.11) | -- | 0.03 (-0.06, 0.11) | -- |
| Age 17 mo | 0.01 (-0.08, 0.10) | 0.58 | 0.01 (-0.05, 0.08) | 0.35 | 0.02 (-0.05, 0.10) | 0.62 | 0.02 (-0.05, 0.09) | 0.95 | 0.01 (-0.07, 0.09) | 0.77 |
| Age 22 mo | -0.05 (-0.13, 0.4) | 0.75 | -0.05 (-0.12, 0.02) | 0.72 | -0.00 (-0.08, 0.08) | 0.94 | 0.04 (-0.04, 0.13) | 0.71 | 0.05 (-0.03, 0.13) | 0.68 |
| Female | -0.04 (-0.11, 0.03) | -- | -0.02 (-0.08, 0.04) | -- | -0.00 (-0.08, 0.07) | -- | 0.03 (-0.04, 0.10) | -- | 0.02 (-0.06, 0.09) | -- |
| Male | -0.01 (-0.08, 0.07) | 0.50 | -0.02 (-0.09, 0.04) | 0.99 | 0.01 (-0.05, 0.07) | 0.72 | 0.02 (-0.06, 0.10) | 0.80 | 0.04 (-0.02, 0.10) | 0.67 |
| Used antibiotics ≥2 times | | | | | | | | | | |
| All obs | 0.00 (-0.03, 0.04) | 0.81 | -0.02 (-0.05, 0.01) | 0.30 | -0.00 (-0.03, 0.03) | 0.84 | -0.01 (-0.05, 0.03) | 0.73 | 0.01 (-0.02, 0.05) | 0.49 |
| Age 6 mo | -0.03 (-0.08, 0.02) | -- | -0.03 (-0.08, 0.03) | -- | -0.02 (-0.08, 0.04) | -- | 0.01 (-0.06, 0.07) | -- | 0.00 (-0.06, 0.07) | -- |
| Age 17 mo | 0.02 (-0.04, 0.08) | 0.20 | 0.00 (-0.05, 0.06) | 0.48 | 0.02 (-0.03, 0.07) | 0.31 | -0.00 (-0.07, 0.06) | 0.85 | 0.02 (-0.04, 0.07) | 0.68 |
| Age 22 mo | 0.02 (-0.04, 0.07) | 0.17 * | -0.03 (-0.07, 0.01) | 0.86 | -0.01 (-0.06, 0.04) | 0.72 | -0.03 (-0.08, 0.03) | 0.42 | 0.02 (-0.02, 0.06) | 0.55 |
| Female | -0.00 (-0.04, 0.04) | -- | -0.00 (-0.04, 0.04) | -- | -0.00 (-0.05, 0.04) | -- | -0.00 (-0.04, 0.04) | -- | -0.00 (-0.05, 0.04) | -- |
| Male | 0.01 (-0.04, 0.07) | 0.69 | -0.03 (-0.08, 0.01) | 0.32 | -0.00 (-0.04, 0,04) | 0.91 | -0.01 (-0.08, 0.05) | 0.81 | 0.03 (-0.02, 0.08) | 0.28 |
| | CD (95% CI) | p-value[a] | CD (95% CI) | p-value[a] | CD (95% CI) | p-value[a] | CD (95% CI) | p-value[a] | CD (95% CI) | p-value[a] |
| Episodes of antibiotic use | | | | | | | | | | |
| All obs | -0.01 (-0.09, 0.07) | 0.77 | -0.03 (-0.10, 0.05) | 0.44 | 0.01 (-0.06, 0.09) | 0.67 | 0.03 (-0.06, 0.12) | 0.56 | 0.05 (-0.05, 0.14) | 0.34 |
| Age 6 mo | -0.06 (-0.17, 0.05) | -- | -0.04 (-0.18, 0.10) | -- | -0.03 (-0.17, 0.12) | -- | 0.03 (-0.13, 0.19) | -- | 0.01 (-0.14, 0.17) | -- |
| Age 17 mo | 0.04 (-0.10, 0.17) | 0.31 | 0.03 (-0.08, 0.13) | 0.41 | 0.07 (-0.04, 0.18) | 0.32 | 0.03 (-0.10, 0.16) | 1.00 | 0.04 (-0.08, 0.16) | 0.78 |
| Age 22 mo | -0.02 (-0.15, 0.10) | 0.67 | -0.08 (-0.18, 0.01) | 0.57 | -0.01 (-0.11, 0.10) | 0.82 | 0.02 (-0.11, 0.15) | 0.89 | 0.08 (-0.03, 0.18) | 0.45 |
| Female | -0.02 (-0.12, 0.07) | -- | -0.01 (-0.10, 0.08) | -- | 0.01 (-0.10, 0.12) | -- | 0.04 (-0.07, 0.14) | -- | 0.02 (-0.10, 0.14) | -- |
| Male | 0.01 (-0.11, 0.12) | 0.64 | -0.05 (-0.16, 0.06) | 0.53 | 0.03 (-0.07, 0.12) | 0.83 | 0.02 (-0.12, 0.16) | 0.85 | 0.07 (-0.03, 0.18) | 0.43 |
| Total days of antibiotic use | | | | | | | | | | |
| All obs | 0.19 (-1.37, 1.76) | 0.81 | -0.59 (-1.61, 0.43) | 0.25 | -0.24 (-1.08, 0.60) | 0.58 | -0.43 (-1.81, 0.94) | 0.53 | 0.34 (-0.48, 1.18) | 0.41 |
| Age 6 mo | -1.55 (-4.27, 1.17) | -- | **-2.71 (-4.88, -0.53)** | -- | -1.67 (-4.16, 0.81) | -- | -0.13 (-1.91, 1.66) | -- | 1.03 (-0.20, 2.26) | -- |
| Age 17 mo | 1.95 (-1.47, 5.37) | 0.11 * | 0.73 (-1.08, 2.54) | 0.02 * | 0.14 (-1.03, 1.31) | 0.26 | -1.81 (-5.22, 1.60) | 0.37 | -0.59 (-2.30, 1.12) | 0.15 * |
| Age 22 mo | 0.00 (-0.72, 0.73) | 0.27 | 0.04 (-0.84, 0.93) | 0.03 * | 0.66 (-0.35, 1.67) | 0.09 * | 0.66 (-0.32, 1.63) | 0.47 | 0.62 (0.46, 1.70) | 0.58 |
| Female | 0.43 (-1.79, 2.64) | -- | -0.51 (-1.84, 0.82) | -- | -0.50 (-1.72, 0.72) | -- | -0.93 (-2.92, 1.06) | -- | 0.01 (-1.16, 1.18) | -- |
| Male | -0.12 (-1.50, 1.27) | 0.61 | -0.65 (-1.99, 0.69) | 0.88 | 0.04 (-1.18, 1.26) | 0.54 | 0.16 (-1.11, 1.42) | 0.29 | 0.69 (-0.47, 1.85) | 0.41 |

PD: Prevalence difference (for binary outcomes); CD: Count difference (for count outcomes); CI: Confidence interval; C: Control; WSH: Water, sanitation, hygiene; N: Nutrition, N+WSH: Nutrition+ water, sanitation, hygiene.

Bolded values refer to estimates with a p-value ≤0.05.

[a] Interaction p-value between study arm and subgroup variable.

* Interaction p-values <0.20 indicate significant effect modification.

**Table S16.** Sensitivity analysis, unadjusted intervention effects on antibiotic use within last 2 weeks and last month, Bangladesh*

| | | | Compared to C | | Compared to WSH | | Compared to N | |
|---|---|---|---|---|---|---|---|---|
| Arm | N | % (n) | PR (95% CI) | p-value | PR (95% CI) | p-value | PR (95% CI) | p-value |
| Bangladesh | | | | | | | | |
| Used antibiotics ≥1 time in last two weeks | | | | | | | | |
| C | 1000 | 21.5 (215) | -- | -- | -- | -- | -- | -- |
| WSH | 1076 | 22.1 (238) | 1.03 (0.84, 1.26) | 0.79 | -- | -- | -- | -- |
| N | 1014 | 19.9 (202) | 0.93 (0.77, 1.11) | 0.42 | -- | -- | -- | -- |
| N+WSH | 1050 | 19.2 (202) | 0.89 (0.73, 1.10) | 0.29 | 0.87 (0.72, 1.06) | 0.16 | 0.97 (0.78, 1.19) | 0.74 |
| Used antibiotics ≥1 time in last month | | | | | | | | |
| C | 1000 | 48.2 (482) | -- | -- | -- | -- | -- | -- |
| WSH | 1076 | 43.2 (465) | 0.90 (0.81, 0.99) | 0.03 | -- | -- | -- | -- |
| N | 1014 | 40.4 (410) | 0.84 (0.76, 0.93) | <0.001 | -- | -- | -- | -- |
| N+WSH | 1050 | 42.3 (444) | 0.88 (0.79, 0.97) | 0.01 | 0.98 (0.88, 1.08) | 0.67 | 1.05 (0.93, 1.18) | 0.45 |

PR: Prevalence ratio; CI: Confidence interval; C: Control; WSH: Water, sanitation, hygiene; N: Nutrition, N+WSH: Nutrition+ water, sanitation, hygiene.

* Not conducted for Kenya because the survey question for this variable in Kenya was designed to also capture non-antibiotic medications.

**Figure S1.** CONSORT Diagram for the WASH Benefits Bangladesh environmental enteric dysfunction substudy

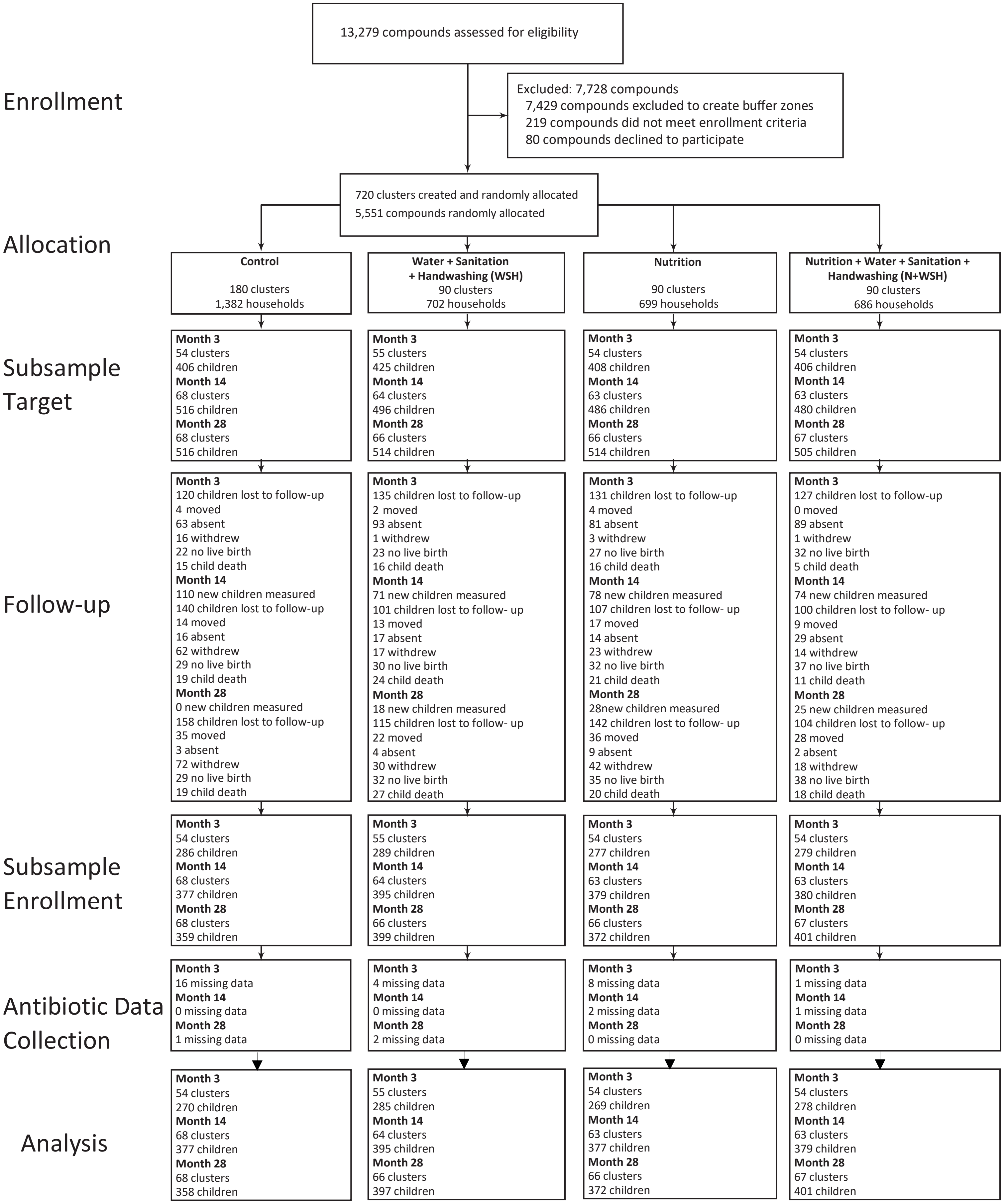

**Figure S2.** CONSORT Diagram for the WASH Benefits Kenya environmental enteric dysfunction substudy

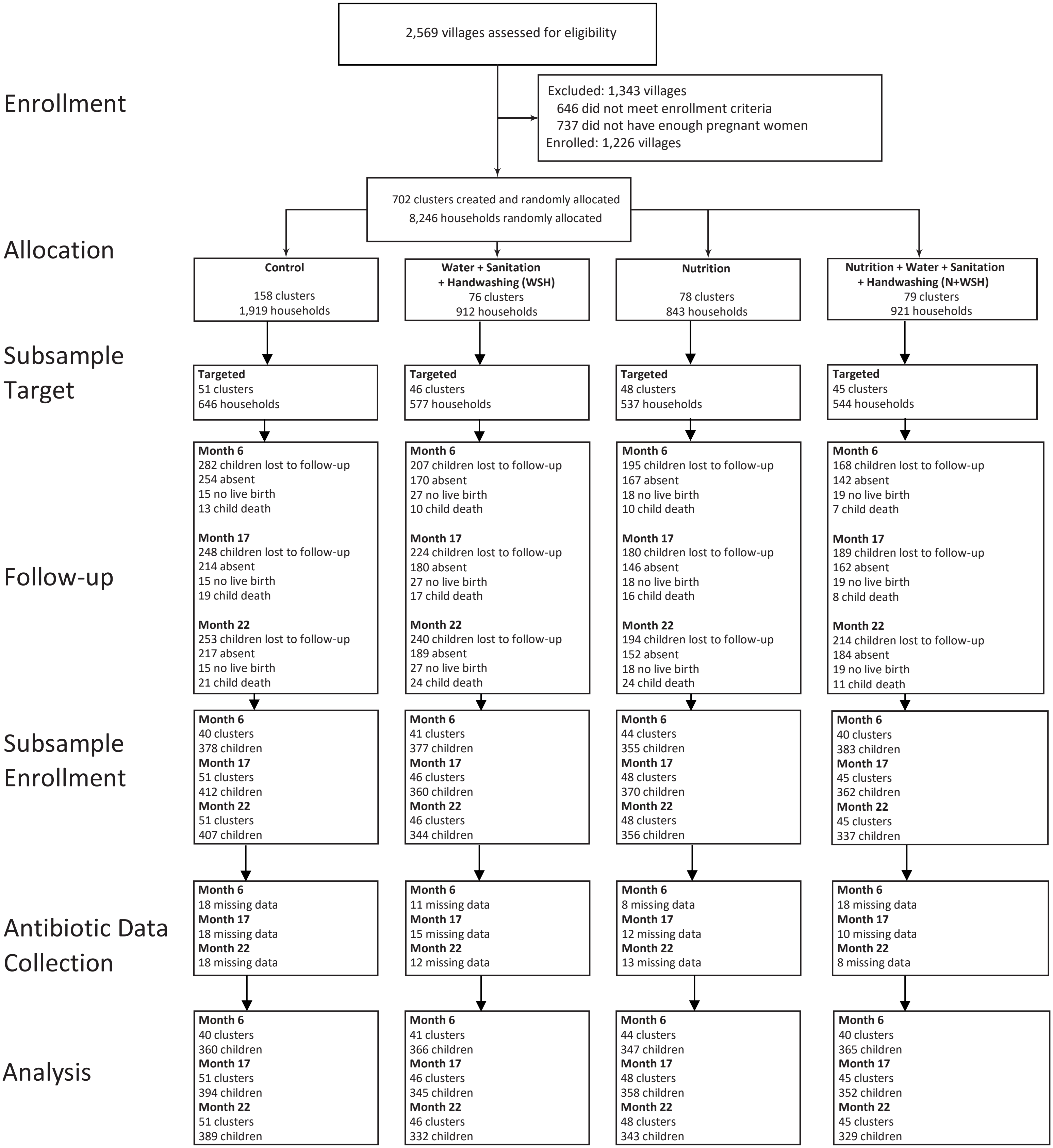
